# Mental distress and health-related quality of life among somatic inpatients: social gradients and hospital mental health implications based on the SomPsyNet study population

**DOI:** 10.1101/2025.11.28.25335384

**Authors:** Gunther Meinlschmidt, Alexander Frick, Iris Baenteli, Christina Karpf, Anja Studer, Sanaa Bahmane, David Buechel, Lukas Ebner, Vincent Ochs, Meike Weber, Marco Bachmann, Andreas Dörner, Sibil Tschudin, Sarah Trost, Kaspar Wyss, Günther Fink, Matthias Schwenkglenks, Rainer Schaefert

## Abstract

**Purpose:** Proactive integrated consultation-liaison psychosomatics and psychiatry in somatic hospitals may improve patients’ mental health beyond inpatient stay. The «SomPsyNet» project targets SOMatic inpatients and aims to prevent PSYchosocial distress by establishing a structured care NETwork based on a Stepped and Collaborative Care Model (SCCM). We used a hospital-based epidemiological perspective to quantify prevalence and social gradients in mental distress and health-related quality of life (HRQoL).

**Methods:** We enrolled inpatients from three tertiary somatic hospitals, estimating how sociodemographic factors were associated with clinically relevant depressive symptoms, anxiety symptoms, somatic symptom distress, overall mental distress (≥1 measure above cut-off), mental, physical, and generic HRQoL, as well as somatic symptom burden.

**Results:** Out of 3,179 participants, 37% showed mental distress. Mental, physical, and generic HRQoL were substantially impaired, while somatic symptom burden was moderate. Younger age and lower income were associated with higher odds of mental distress, lower mental HRQoL, and greater somatic symptom burden (all *p-for-trend*<0.05), yet younger patients showed better physical HRQoL, and higher income was associated with better generic HRQoL (*p-for-trend*<0.05). Having no Swiss citizenship or having an employment situation affected by disability, accident, or illness, was associated with worse outcomes across most (*p*’s<0.01) or all (*p*’s<0.05) indicators, respectively. Other factors showed less consistent associations.

**Conclusions:** A substantial proportion of somatic inpatients in the hospitals studied experienced mental distress and showed impaired HRQoL. Findings identified priority groups for hospital public mental health and inform resource planning for stepped, collaborative care and culturally/linguistically adapted services in acute somatic settings.

**Statements and Declarations:** *Declaration of Competing Interest & Funding Independent of the Project:* G.M. & R.S. received funding in the context of a Horizon Europe project from the Swiss State Secretariat for Education, Research and Innovation (SERI) under contract number 22.00094. Further, G.M. & R.S. received funding from Wings Health Inc. in the context of a proof-of-concept study. G.M. received funding from the Swiss Heart Foundation under project no. FF21101, from the Research Foundation of the International Psychoanalytic University (IPU) Berlin under projects no. 5087 and 5217, from the Swiss National Science Foundation (SNSF) under project no. 100014_135328, from the German Federal Ministry of Education and Research under budget item 68606, and from the Hasler Foundation under project no. 23004. G.M. is co-founder and holds stock in Therayou AG, which is active in the field of digital and blended mental healthcare. G.M. receives royalties from publishing companies as author, including a book published by Springer, and an honorarium from Lundbeck for speaking at a symposium. Furthermore, G.M. is compensated for providing psychotherapy to patients, acting as a supervisor, serving as a self-experience facilitator (‘*Selbsterfahrungsleiter*’), and for postgraduate training of psychotherapists and supervisors.

R.S. is co-editor of the German AWMF S3-Guidelines on Functional Complaints, and contributed to the German guidelines on irritable bowel syndrome, and on Lyme Borreliosis. R.S. is chairman of the Basel Institute for Psychosomatic Medicine (BIPM) and founder and managing director of the Psychosomatic and Psychosocial Services GmbH, that develops and implements psychosomatic and psychosocial training and continuing education programs. The authors declare no other potential conflict of interests. The research activities were fully independent and there were no intellectual or financial proprietary claims.

## 1. Introduction

Mental health conditions contribute substantially to population morbidity and costs [1, 2]. In Europe, around 38% of adults experience clinically relevant mental disorders annually [3–5]. Beyond diagnosed mental disorders, psychosocial distress – common in somatic illness – impairs functioning and quality of life [6, 7].

In somatic hospitals, mental–somatic multimorbidity is frequent and costly: clinically relevant depression, anxiety, and somatic symptom distress are linked to longer stays and readmissions, yet often remain underdetected [8, 9]. This exacerbates resource utilization and healthcare costs [10, 11]. Longitudinal data further indicate mental–physical interplay [12], supporting integrated approaches in routine care [7, 13]. Consistently, a meta-analysis of consultation-liaison (CL) services in general hospitals reported small-to-moderate symptom improvements for depression and anxiety compared with usual care [14]. Relatedly, in acute medical contexts, comorbid anxiety can shape time-critical care pathways; for instance, the multicentre MEDEA study found generalized anxiety disorder to be associated with altered prehospital delay patterns in acute myocardial infarction [15].

Health policies such as the Swiss National Strategy on NCD Prevention (2017–2028) advocate for addressing psychosocial factors together with somatic conditions. Collaborative and integrated care models improve mental health outcomes in medical settings [16–19]. In a German cluster-randomised trial, guideline-based stepped and collaborative care produced clinically meaningful improvements in depression versus usual care [20]. “Proactive integrated consultation-liaison psychiatry (PICLP)” models show benefits for older inpatients and hospital operations [21].

SomPsyNet developed a Stepped and Collaborative Care Model (SCCM) [22], to proactively identify and address psychosocial distress among somatic inpatients [23–26].

Here we report baseline (study-entry) mental health status of somatic hospital inpatients based on data from the SomPsyNet trial, (1) characterizing the prevalence and intensity of different forms of mental distress, HRQoL (mental, physical, as well as generic), and somatic symptom burden; and (2) examining how these outcomes vary by patient sociodemographics, to guide targeted stepped, collaborative interventions [27].

## 2. Methods

### 2.1. Study setting and stepped-wedge cluster randomized design

We conducted a cross-sectional baseline analysis within the SomPsyNet study, a stepped-wedge cluster randomized trial across three tertiary hospitals, being part of the SomPsyNet project that aimed at inpatients from tertiary SOMatic hospitals to promote prevention of PSYchosocial distress by establishing a structured care NETwork (Meinlschmidt et al., 2023). Clusters (wards) transitioned from (i) treatment as usual, to (ii) treatment as usual plus screening, to (iii) full SCCM with intervention recommendations for positive screens. For this cross-sectional analysis, we used study entry (baseline) assessments obtained across study-phases to characterize mental distress and HRQoL at admission.

### 2.2. Recruitment, inclusion and exclusion criteria

Recruitment occurred from 09-06-2020 to 16-12-2022 across the UHB, BESP, and UAFP in selected wards. Consecutive admissions were screened via hospital systems and ward liaison; eligibility was verified from records and patient interviews. Eligible patients were informed about the study and had to consent before being included. For patients with multiple admissions, only the first was analyzed. Should a participant choose to withdraw, their data collection ceased and previously collected data were excluded from analyses. Eligible participants were those hospitalized in a participating ward during the study period. Exclusion criteria included: age <18 years, insufficient German proficiency, lack of independent consent capacity; severe medical instability precluding participation; acute suicidality; conditions covered by specific psychosocial services (e.g., oncology; gender-affirming care); previous SomPsyNet participation; active Coronavirus disease 2019 (COVID-19) infection; or medical responsibility by a non-participating ward despite location in a participating ward.

### 2.3. Data collection

Participants completed a baseline questionnaire (tablet-assisted ‘heartbeat one’, Heartbeat Medical Solutions GmbH, Berlin, Germany; paper or staff-assisted if needed). Data were managed in secuTrial®. Sample size targets for the parent stepped-wedge trial (primary outcome: SF-36 MCS) were 2,200–2,500, as detailed elsewhere [22]. To elucidate potential selection bias, we compared the SomPsyNet cohort’s age/sex distribution with ward registry data from the same period (Online Resource 1).

### 2.4. Study variables

The full set of data collected has been described elsewhere [22]. Sociodemographic characteristics included age, gender, nationality, household income, marital status, living arrangement, education, and employment (questionnaire/hospital records).

Mental distress was assessed along three indicators, using for depressive symptoms the 8-item Patient Health Questionnaire (PHQ-8) [28], for anxiety symptoms the Generalized Anxiety Disorder-7 scale (GAD-7) [29]; and for somatic symptom distress, the Somatic Symptom Disorder–B Criteria Scale (SSD-12) [30], applying validated cut-offs (PHQ-8≥10; GAD-7≥10; SSD-12≥23).

HRQoL comprised mental HRQoL measured by the ‘Mental Health Component Summary (MCS) score’ of the Short Form (36) Health Survey (SF-36) version 1 (primary outcome of the parent trial) [31–33], physical HRQoL measured by the SF-36 Physical Health Component Summary (PCS) score, and generic HRQoL with the European Quality of Life – 5 Dimensions questionnaire – 5 level version (EQ-5D-5L index) [34, 35]. We evaluated Somatic symptom burden using the Somatic Symptom Scale-8 (SSS-8) [36]. SF-36 psychometrics and German translations are well established [31–33]. The validity, reliability, and clinical utility of all selected instruments has been previously established within multiple other studies. Staff were trained in administration and data entry.

### 2.5. Handling of quantitative variables

Sociodemographic categories followed Online Resource 2. We addressed Item-level missingness per published rules [37, 38]: PHQ-8/GAD-7 – if ≥6 items completed, replace missing with mean of completed items; SSD-12 – if ≥9 items completed, use mean of completed items; SF-36 MCS/PCS – procedure provided by RAND 36-Item Health Survey (Version 1.0) [31], using the mean of the completed items. Following standard instructions previously described [39], we applied the SF-36 normative data that are based on a Swiss representative sample, provided by Roser et al. [40]. Roser et al. [40] used the SF-36 version 2 while we used version 1. Yet, the two versions do only slightly differ from each other [41] (Kroenke et al., 2009). We scored EQ-5D-5L only if all five dimensions were complete [42, 43]. For SSS-8, if ≥6 of 8 items were completed, missing items were replaced by the mean of completed items; otherwise, the scale was set to missing [36]. If participant did not indicate to be of Swiss citizenship, we counted them as non-Swiss citizenship. Scale computation followed instrument manuals and publications [29, 39, 41, 44].

### 2.6. Patient and public involvement

A patient participation committee contributed to grant preparation, study design, consent materials, recruitment strategies, and dissemination. Patient representatives advised on interfaces between sectors and the enrolment process.

### 2.7. Statistical methods

Analytic details are expanded in the Online Resource 3. We summarized sample characteristics and outcomes with appropriate descriptive statistics and provided 95% Confidence Intervals (CIs) where appropriate.

We simultaneously entered all sociodemographic predictors (see Table 1) in separate logistic or generalized linear models for each outcome (above PHQ-8, GAD-7, SSD-12 cutoffs or any of these; MCS, PCS, SSS-8, EQ-5D-5L), followed by p-for-trend analyses in case of ordinal predictors. We used generalized linear models to accommodate heteroskedasticity and non-normality; results are reported on original units. We handled missing data via multiple imputation by chained equations (m = 100 imputed datasets, with estimates pooled across imputations).

**Table 1.**
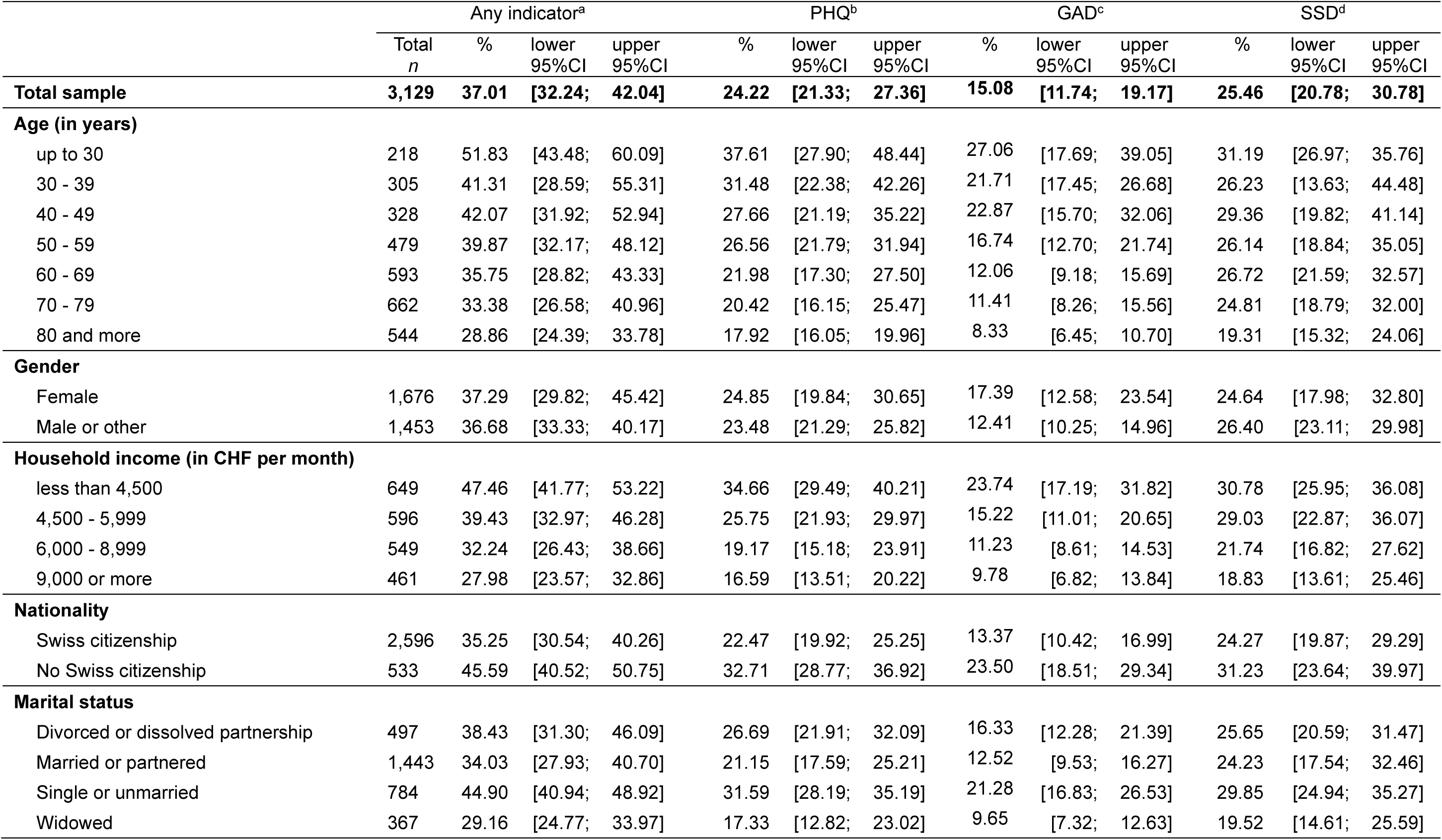
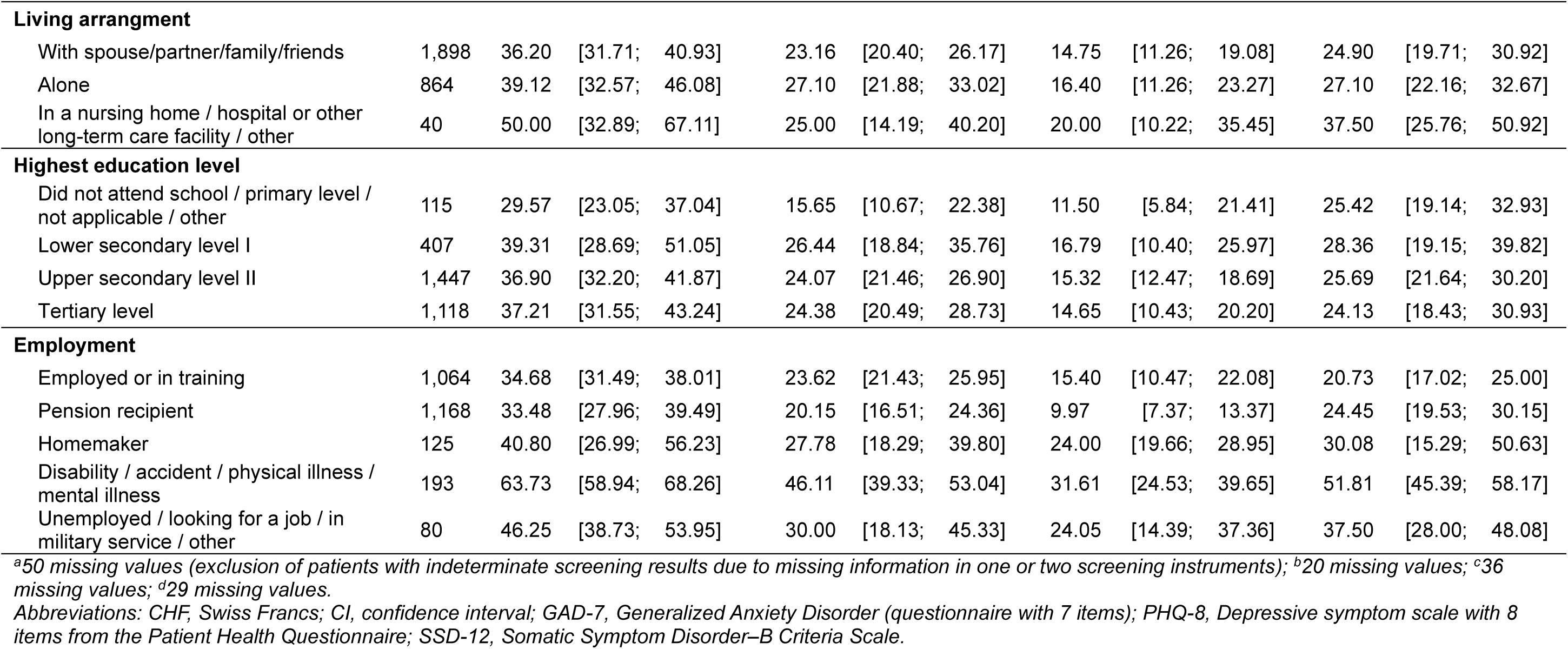
Patients scoring above cut-off in mental health screening instruments.

|  | Any indicator<sup>a</sup> |  |  |  | PHQ<sup>b</sup> |  |  | GAD<sup>c</sup> |  |  | SSD<sup>d</sup> |  |  |
| --- | --- | --- | --- | --- | --- | --- | --- | --- | --- | --- | --- | --- | --- |
|  | Total<br>*n* | % | lower<br>95%CI | upper<br>95%CI | % | lower<br>95%CI | upper<br>95%CI | % | lower<br>95%CI | upper<br>95%CI | % | lower<br>95%CI | upper<br>95%CI |
| **Total sample** | **3,129** | **37.01** | **[32.24;** | **42.04]** | **24.22** | **[21.33;** | **27.36]** | **15.08** | **[11.74;** | **19.17]** | **25.46** | **[20.78;** | **30.78]** |
| **Age (in years)** |  |  |  |  |  |  |  |  |  |  |  |  |  |
| up to 30 | 218 | 51.83 | [43.48; | 60.09] | 37.61 | [27.90; | 48.44] | 27.06 | [17.69; | 39.05] | 31.19 | [26.97; | 35.76] |
| 30 - 39 | 305 | 41.31 | [28.59; | 55.31] | 31.48 | [22.38; | 42.26] | 21.71 | [17.45; | 26.68] | 26.23 | [13.63; | 44.48] |
| 40 - 49 | 328 | 42.07 | [31.92; | 52.94] | 27.66 | [21.19; | 35.22] | 22.87 | [15.70; | 32.06] | 29.36 | [19.82; | 41.14] |
| 50 - 59 | 479 | 39.87 | [32.17; | 48.12] | 26.56 | [21.79; | 31.94] | 16.74 | [12.70; | 21.74] | 26.14 | [18.84; | 35.05] |
| 60 - 69 | 593 | 35.75 | [28.82; | 43.33] | 21.98 | [17.30; | 27.50] | 12.06 | [9.18; | 15.69] | 26.72 | [21.59; | 32.57] |
| 70 - 79 | 662 | 33.38 | [26.58; | 40.96] | 20.42 | [16.15; | 25.47] | 11.41 | [8.26; | 15.56] | 24.81 | [18.79; | 32.00] |
| 80 and more | 544 | 28.86 | [24.39; | 33.78] | 17.92 | [16.05; | 19.96] | 8.33 | [6.45; | 10.70] | 19.31 | [15.32; | 24.06] |
| **Gender** |  |  |  |  |  |  |  |  |  |  |  |  |  |
| Female | 1,676 | 37.29 | [29.82; | 45.42] | 24.85 | [19.84; | 30.65] | 17.39 | [12.58; | 23.54] | 24.64 | [17.98; | 32.80] |
| Male or other | 1,453 | 36.68 | [33.33; | 40.17] | 23.48 | [21.29; | 25.82] | 12.41 | [10.25; | 14.96] | 26.40 | [23.11; | 29.98] |
| **Household income (in CHF per month)** |  |  |  |  |  |  |  |  |  |  |  |  |  |
| less than 4,500 | 649 | 47.46 | [41.77; | 53.22] | 34.66 | [29.49; | 40.21] | 23.74 | [17.19; | 31.82] | 30.78 | [25.95; | 36.08] |
| 4,500 - 5,999 | 596 | 39.43 | [32.97; | 46.28] | 25.75 | [21.93; | 29.97] | 15.22 | [11.01; | 20.65] | 29.03 | [22.87; | 36.07] |
| 6,000 - 8,999 | 549 | 32.24 | [26.43; | 38.66] | 19.17 | [15.18; | 23.91] | 11.23 | [8.61; | 14.53] | 21.74 | [16.82; | 27.62] |
| 9,000 or more | 461 | 27.98 | [23.57; | 32.86] | 16.59 | [13.51; | 20.22] | 9.78 | [6.82; | 13.84] | 18.83 | [13.61; | 25.46] |
| **Nationality** |  |  |  |  |  |  |  |  |  |  |  |  |  |
| Swiss citizenship | 2,596 | 35.25 | [30.54; | 40.26] | 22.47 | [19.92; | 25.25] | 13.37 | [10.42; | 16.99] | 24.27 | [19.87; | 29.29] |
| No Swiss citizenship | 533 | 45.59 | [40.52; | 50.75] | 32.71 | [28.77; | 36.92] | 23.50 | [18.51; | 29.34] | 31.23 | [23.64; | 39.97] |
| **Marital status** |  |  |  |  |  |  |  |  |  |  |  |  |  |
| Divorced or dissolved partnership | 497 | 38.43 | [31.30; | 46.09] | 26.69 | [21.91; | 32.09] | 16.33 | [12.28; | 21.39] | 25.65 | [20.59; | 31.47] |
| Married or partnered | 1,443 | 34.03 | [27.93; | 40.70] | 21.15 | [17.59; | 25.21] | 12.52 | [9.53; | 16.27] | 24.23 | [17.54; | 32.46] |
| Single or unmarried | 784 | 44.90 | [40.94; | 48.92] | 31.59 | [28.19; | 35.19] | 21.28 | [16.83; | 26.53] | 29.85 | [24.94; | 35.27] |
| Widowed | 367 | 29.16 | [24.77; | 33.97] | 17.33 | [12.82; | 23.02] | 9.65 | [7.32; | 12.63] | 19.52 | [14.61; | 25.59] |

|  |  |  |  |  |  |  |  |  |  |
| --- | --- | --- | --- | --- | --- | --- | --- | --- | --- |
| **Living arrangement** |  |  |  |  |  |  |  |  |  |
| With spouse/partner/family/friends | 1,898 | 36.20 | [31.71; 40.93] | 23.16 | [20.40; 26.17] | 14.75 | [11.26; 19.08] | 24.90 | [19.71; 30.92] |
| Alone | 864 | 39.12 | [32.57; 46.08] | 27.10 | [21.88; 33.02] | 16.40 | [11.26; 23.27] | 27.10 | [22.16; 32.67] |
| In a nursing home / hospital or other long-term care facility / other | 40 | 50.00 | [32.89; 67.11] | 25.00 | [14.19; 40.20] | 20.00 | [10.22; 35.45] | 37.50 | [25.76; 50.92] |
| **Highest education level** |  |  |  |  |  |  |  |  |  |
| Did not attend school / primary level / not applicable / other | 115 | 29.57 | [23.05; 37.04] | 15.65 | [10.67; 22.38] | 11.50 | [5.84; 21.41] | 25.42 | [19.14; 32.93] |
| Lower secondary level I | 407 | 39.31 | [28.69; 51.05] | 26.44 | [18.84; 35.76] | 16.79 | [10.40; 25.97] | 28.36 | [19.15; 39.82] |
| Upper secondary level II | 1,447 | 36.90 | [32.20; 41.87] | 24.07 | [21.46; 26.90] | 15.32 | [12.47; 18.69] | 25.69 | [21.64; 30.20] |
| Tertiary level | 1,118 | 37.21 | [31.55; 43.24] | 24.38 | [20.49; 28.73] | 14.65 | [10.43; 20.20] | 24.13 | [18.43; 30.93] |
| **Employment** |  |  |  |  |  |  |  |  |  |
| Employed or in training | 1,064 | 34.68 | [31.49; 38.01] | 23.62 | [21.43; 25.95] | 15.40 | [10.47; 22.08] | 20.73 | [17.02; 25.00] |
| Pension recipient | 1,168 | 33.48 | [27.96; 39.49] | 20.15 | [16.51; 24.36] | 9.97 | [7.37; 13.37] | 24.45 | [19.53; 30.15] |
| Homemaker | 125 | 40.80 | [26.99; 56.23] | 27.78 | [18.29; 39.80] | 24.00 | [19.66; 28.95] | 30.08 | [15.29; 50.63] |
| Disability / accident / physical illness / mental illness | 193 | 63.73 | [58.94; 68.26] | 46.11 | [39.33; 53.04] | 31.61 | [24.53; 39.65] | 51.81 | [45.39; 58.17] |
| Unemployed / looking for a job / in military service / other | 80 | 46.25 | [38.73; 53.95] | 30.00 | [18.13; 45.33] | 24.05 | [14.39; 37.36] | 37.50 | [28.00; 48.08] |

<sup>a</sup>50 missing values (exclusion of patients with indeterminate screening results due to missing information in one or two screening instruments); <sup>b</sup>20 missing values; <sup>c</sup>36 missing values; <sup>d</sup>29 missing values.
Abbreviations: CHF, Swiss Francs; CI, confidence interval; GAD-7, Generalized Anxiety Disorder (questionnaire with 7 items); PHQ-8, Depressive symptom scale with 8 items from the Patient Health Questionnaire; SSD-12, Somatic Symptom Disorder–B Criteria Scale.

To account for the sampling design and ensure robust variance estimation, we used Stata svy with Taylor series linearization, clustering patients by a nine-level variable (reflecting 7 triplets with specialty differences).

All regression analyses accounted for the clustered study design. Analyses used Stata’s svy commands with Taylor series linearization. Collinearity diagnostics (Spearman’s rho and variance inflation factors, VIFs) showed no problematic multicollinearity (all VIFs < 5).

## 3. Results

A flowchart detailing the inclusion of study subjects is provided in Online Resource 4.

### 3.1. Sample Characteristics and Stratified Screening, Clinical and HRQoL Outcomes

Our analyses encompassed data from a total of 3,179 somatic hospital inpatients. We present sociodemographic characteristics of the total study population in Online Resource 2, frequencies of patients scoring above cut-off in the mental health screening instruments by sociodemographic factors in Table 1, and descriptives of the continuous variables MCS, PCS, EQ-5D-5L, and SSS-8 stratified by sociodemographic factors in Table 2. Comparing the gender and age distributions of the SomPsyNet study participants with hospital registry data from the same wards and time period indicated possible selective participation, revealing an approximately 4% higher proportion of males (46.2% vs. 41.9%) than would have been expected, as well as more younger (i.e., below 70 years) and fewer older (i.e., 70 years and more) participants than would have been expected if the SomPsyNet sample were entirely representative of all patients hospitalized at the same wards during the same time period (see Online Resource 1).

**Table 2.**
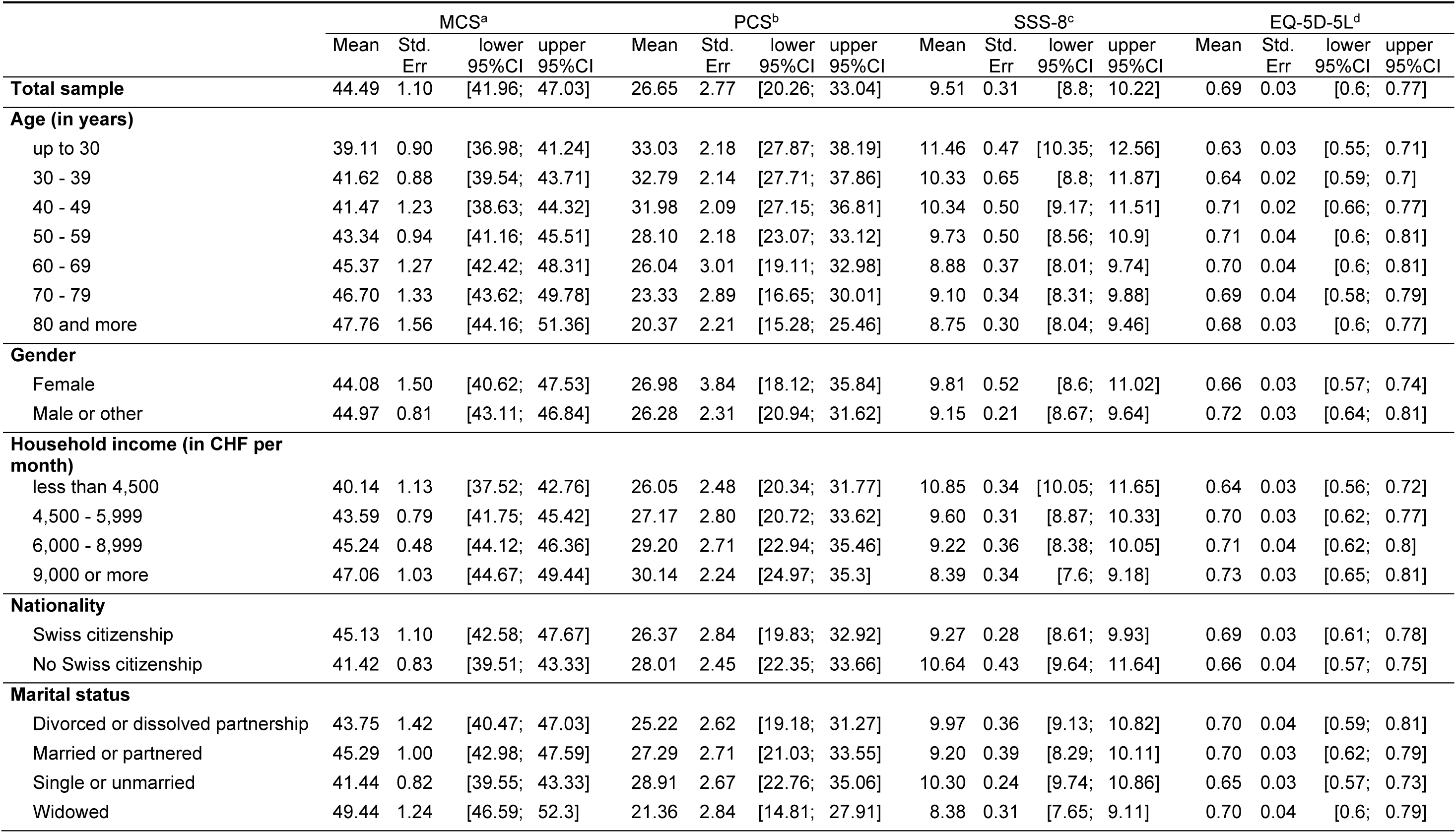
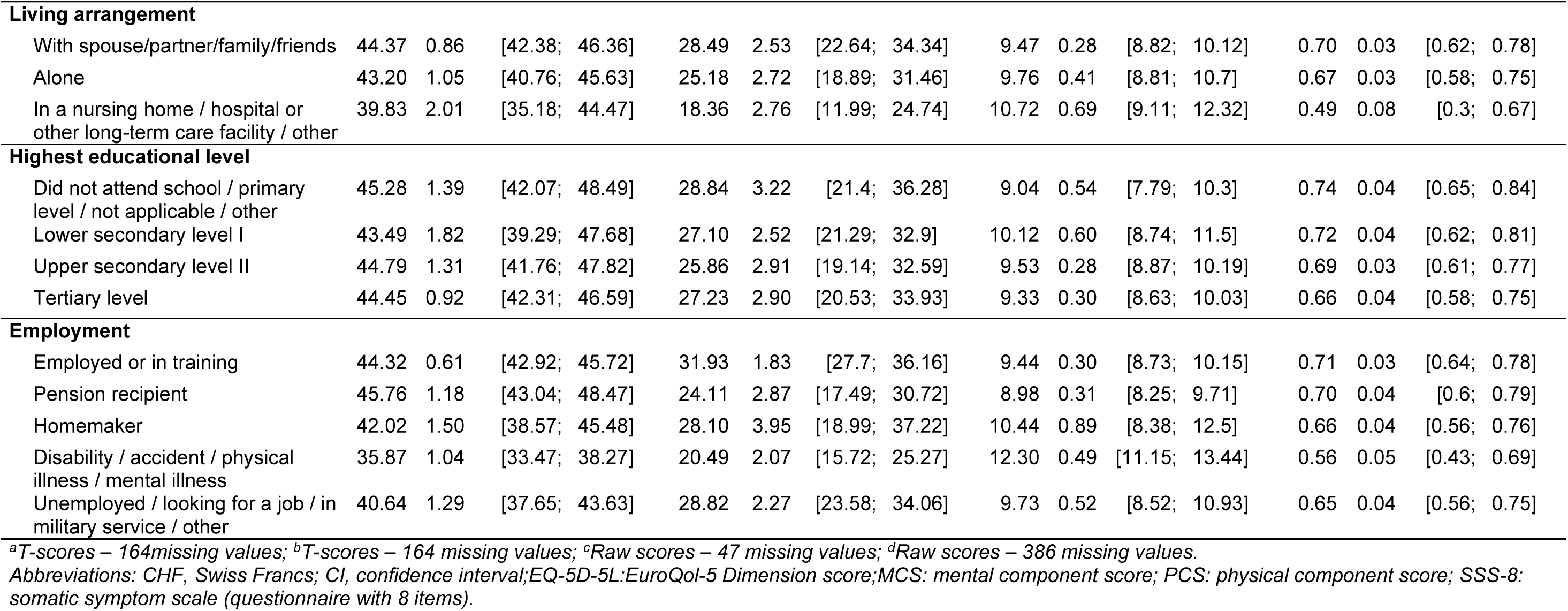
Descriptives of the continuous variables MCS, PCS, SSS-8, and EQ-5D-5L, stratified by sociodemographic factors.

|  | MCS<sup>a</sup> |  |  |  | PCS<sup>b</sup> |  |  |  | SSS-8<sup>c</sup> |  |  |  | EQ-5D-5L<sup>d</sup> |  |  |  |
| --- | --- | --- | --- | --- | --- | --- | --- | --- | --- | --- | --- | --- | --- | --- | --- | --- |
|  | Mean | Std. Err | lower 95%CI | upper 95%CI | Mean | Std. Err | lower 95%CI | upper 95%CI | Mean | Std. Err | lower 95%CI | upper 95%CI | Mean | Std. Err | lower 95%CI | upper 95%CI |
| **Total sample** | 44.49 | 1.10 | [41.96; 47.03] |  | 26.65 | 2.77 | [20.26; 33.04] |  | 9.51 | 0.31 | [8.8; 10.22] |  | 0.69 | 0.03 | [0.6; 0.77] |  |
| **Age (in years)** |  |  |  |  |  |  |  |  |  |  |  |  |  |  |  |  |
| up to 30 | 39.11 | 0.90 | [36.98; 41.24] |  | 33.03 | 2.18 | [27.87; 38.19] |  | 11.46 | 0.47 | [10.35; 12.56] |  | 0.63 | 0.03 | [0.55; 0.71] |  |
| 30 - 39 | 41.62 | 0.88 | [39.54; 43.71] |  | 32.79 | 2.14 | [27.71; 37.86] |  | 10.33 | 0.65 | [8.8; 11.87] |  | 0.64 | 0.02 | [0.59; 0.7] |  |
| 40 - 49 | 41.47 | 1.23 | [38.63; 44.32] |  | 31.98 | 2.09 | [27.15; 36.81] |  | 10.34 | 0.50 | [9.17; 11.51] |  | 0.71 | 0.02 | [0.66; 0.77] |  |
| 50 - 59 | 43.34 | 0.94 | [41.16; 45.51] |  | 28.10 | 2.18 | [23.07; 33.12] |  | 9.73 | 0.50 | [8.56; 10.9] |  | 0.71 | 0.04 | [0.6; 0.81] |  |
| 60 - 69 | 45.37 | 1.27 | [42.42; 48.31] |  | 26.04 | 3.01 | [19.11; 32.98] |  | 8.88 | 0.37 | [8.01; 9.74] |  | 0.70 | 0.04 | [0.6; 0.81] |  |
| 70 - 79 | 46.70 | 1.33 | [43.62; 49.78] |  | 23.33 | 2.89 | [16.65; 30.01] |  | 9.10 | 0.34 | [8.31; 9.88] |  | 0.69 | 0.04 | [0.58; 0.79] |  |
| 80 and more | 47.76 | 1.56 | [44.16; 51.36] |  | 20.37 | 2.21 | [15.28; 25.46] |  | 8.75 | 0.30 | [8.04; 9.46] |  | 0.68 | 0.03 | [0.6; 0.77] |  |
| **Gender** |  |  |  |  |  |  |  |  |  |  |  |  |  |  |  |  |
| Female | 44.08 | 1.50 | [40.62; 47.53] |  | 26.98 | 3.84 | [18.12; 35.84] |  | 9.81 | 0.52 | [8.6; 11.02] |  | 0.66 | 0.03 | [0.57; 0.74] |  |
| Male or other | 44.97 | 0.81 | [43.11; 46.84] |  | 26.28 | 2.31 | [20.94; 31.62] |  | 9.15 | 0.21 | [8.67; 9.64] |  | 0.72 | 0.03 | [0.64; 0.81] |  |
| **Household income (in CHF per month)** |  |  |  |  |  |  |  |  |  |  |  |  |  |  |  |  |
| less than 4,500 | 40.14 | 1.13 | [37.52; 42.76] |  | 26.05 | 2.48 | [20.34; 31.77] |  | 10.85 | 0.34 | [10.05; 11.65] |  | 0.64 | 0.03 | [0.56; 0.72] |  |
| 4,500 - 5,999 | 43.59 | 0.79 | [41.75; 45.42] |  | 27.17 | 2.80 | [20.72; 33.62] |  | 9.60 | 0.31 | [8.87; 10.33] |  | 0.70 | 0.03 | [0.62; 0.77] |  |
| 6,000 - 8,999 | 45.24 | 0.48 | [44.12; 46.36] |  | 29.20 | 2.71 | [22.94; 35.46] |  | 9.22 | 0.36 | [8.38; 10.05] |  | 0.71 | 0.04 | [0.62; 0.8] |  |
| 9,000 or more | 47.06 | 1.03 | [44.67; 49.44] |  | 30.14 | 2.24 | [24.97; 35.3] |  | 8.39 | 0.34 | [7.6; 9.18] |  | 0.73 | 0.03 | [0.65; 0.81] |  |
| **Nationality** |  |  |  |  |  |  |  |  |  |  |  |  |  |  |  |  |
| Swiss citizenship | 45.13 | 1.10 | [42.58; 47.67] |  | 26.37 | 2.84 | [19.83; 32.92] |  | 9.27 | 0.28 | [8.61; 9.93] |  | 0.69 | 0.03 | [0.61; 0.78] |  |
| No Swiss citizenship | 41.42 | 0.83 | [39.51; 43.33] |  | 28.01 | 2.45 | [22.35; 33.66] |  | 10.64 | 0.43 | [9.64; 11.64] |  | 0.66 | 0.04 | [0.57; 0.75] |  |
| **Marital status** |  |  |  |  |  |  |  |  |  |  |  |  |  |  |  |  |
| Divorced or dissolved partnership | 43.75 | 1.42 | [40.47; 47.03] |  | 25.22 | 2.62 | [19.18; 31.27] |  | 9.97 | 0.36 | [9.13; 10.82] |  | 0.70 | 0.04 | [0.59; 0.81] |  |
| Married or partnered | 45.29 | 1.00 | [42.98; 47.59] |  | 27.29 | 2.71 | [21.03; 33.55] |  | 9.20 | 0.39 | [8.29; 10.11] |  | 0.70 | 0.03 | [0.62; 0.79] |  |
| Single or unmarried | 41.44 | 0.82 | [39.55; 43.33] |  | 28.91 | 2.67 | [22.76; 35.06] |  | 10.30 | 0.24 | [9.74; 10.86] |  | 0.65 | 0.03 | [0.57; 0.73] |  |
| Widowed | 49.44 | 1.24 | [46.59; 52.3] |  | 21.36 | 2.84 | [14.81; 27.91] |  | 8.38 | 0.31 | [7.65; 9.11] |  | 0.70 | 0.04 | [0.6; 0.79] |  |

|  |  |  |  |  |  |  |  |  |  |  |  |  |
| --- | --- | --- | --- | --- | --- | --- | --- | --- | --- | --- | --- | --- |
| **Living arrangement** |  |  |  |  |  |  |  |  |  |  |  |  |
| With spouse/partner/family/friends | 44.37 | 0.86 | [42.38; 46.36] | 28.49 | 2.53 | [22.64; 34.34] | 9.47 | 0.28 | [8.82; 10.12] | 0.70 | 0.03 | [0.62; 0.78] |
| Alone | 43.20 | 1.05 | [40.76; 45.63] | 25.18 | 2.72 | [18.89; 31.46] | 9.76 | 0.41 | [8.81; 10.7] | 0.67 | 0.03 | [0.58; 0.75] |
| In a nursing home / hospital or other long-term care facility / other | 39.83 | 2.01 | [35.18; 44.47] | 18.36 | 2.76 | [11.99; 24.74] | 10.72 | 0.69 | [9.11; 12.32] | 0.49 | 0.08 | [0.3; 0.67] |
| **Highest educational level** |  |  |  |  |  |  |  |  |  |  |  |  |
| Did not attend school / primary level / not applicable / other | 45.28 | 1.39 | [42.07; 48.49] | 28.84 | 3.22 | [21.4; 36.28] | 9.04 | 0.54 | [7.79; 10.3] | 0.74 | 0.04 | [0.65; 0.84] |
| Lower secondary level I | 43.49 | 1.82 | [39.29; 47.68] | 27.10 | 2.52 | [21.29; 32.9] | 10.12 | 0.60 | [8.74; 11.5] | 0.72 | 0.04 | [0.62; 0.81] |
| Upper secondary level II | 44.79 | 1.31 | [41.76; 47.82] | 25.86 | 2.91 | [19.14; 32.59] | 9.53 | 0.28 | [8.87; 10.19] | 0.69 | 0.03 | [0.61; 0.77] |
| Tertiary level | 44.45 | 0.92 | [42.31; 46.59] | 27.23 | 2.90 | [20.53; 33.93] | 9.33 | 0.30 | [8.63; 10.03] | 0.66 | 0.04 | [0.58; 0.75] |
| **Employment** |  |  |  |  |  |  |  |  |  |  |  |  |
| Employed or in training | 44.32 | 0.61 | [42.92; 45.72] | 31.93 | 1.83 | [27.7; 36.16] | 9.44 | 0.30 | [8.73; 10.15] | 0.71 | 0.03 | [0.64; 0.78] |
| Pension recipient | 45.76 | 1.18 | [43.04; 48.47] | 24.11 | 2.87 | [17.49; 30.72] | 8.98 | 0.31 | [8.25; 9.71] | 0.70 | 0.04 | [0.6; 0.79] |
| Homemaker | 42.02 | 1.50 | [38.57; 45.48] | 28.10 | 3.95 | [18.99; 37.22] | 10.44 | 0.89 | [8.38; 12.5] | 0.66 | 0.04 | [0.56; 0.76] |
| Disability / accident / physical illness / mental illness | 35.87 | 1.04 | [33.47; 38.27] | 20.49 | 2.07 | [15.72; 25.27] | 12.30 | 0.49 | [11.15; 13.44] | 0.56 | 0.05 | [0.43; 0.69] |
| Unemployed / looking for a job / in military service / other | 40.64 | 1.29 | [37.65; 43.63] | 28.82 | 2.27 | [23.58; 34.06] | 9.73 | 0.52 | [8.52; 10.93] | 0.65 | 0.04 | [0.56; 0.75] |

<sup>a</sup>T-scores – 164 missing values; <sup>b</sup>T-scores – 164 missing values; <sup>c</sup>Raw scores – 47 missing values; <sup>d</sup>Raw scores – 386 missing values.
Abbreviations: CHF, Swiss Francs; CI, confidence interval; EQ-5D-5L: EuroQol-5 Dimension score; MCS: mental component score; PCS: physical component score; SSS-8: somatic symptom scale (questionnaire with 8 items).

Overall, 37.0% (95%CI: 32.2% – 42.1%) of somatic hospital inpatients scored at or above the cut-off on at least one of the mental health screening instruments. Accordingly, 63.0% of patients reported no relevant mental symptoms in the domains assessed. Specifically, 24.2% (95%CI: 21.3% – 27.4%) screened positive for depressive symptoms (PHQ-8 ≥ 10), 15.1% (95%CI: 11.7% – 19.2%) for anxiety (GAD-7 ≥10), and 25.5% (95%CI: 20.7% – 30.8%) for somatic distress (SSD-12 ≥ 23) (see Table 1).

Multiple (2 or 3) areas of mental distress were reported by 20.3% of the population. A Venn diagram illustrating the overlap in mental distress indicators among patients is provided in Figure 1.

**Fig. 1.**
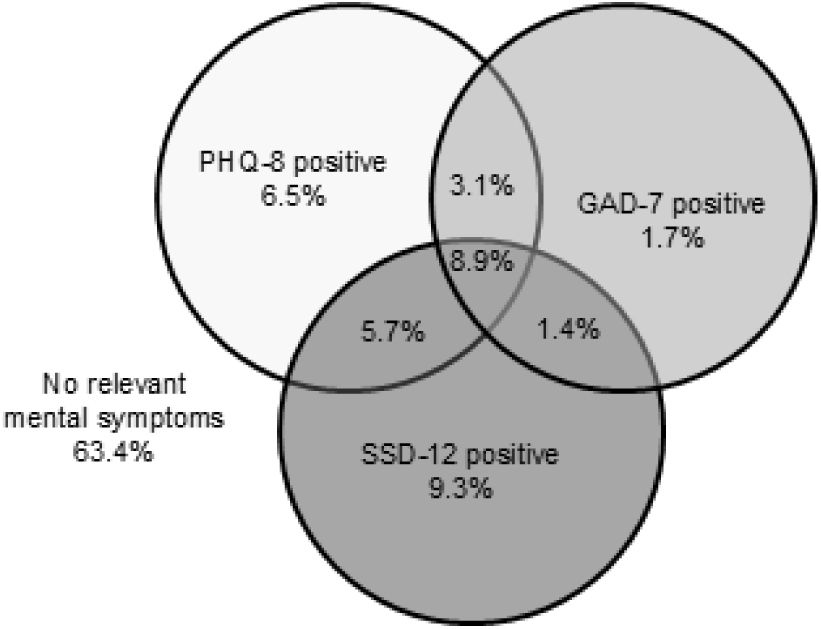
Venn diagram depicting overlap of mental symptoms. Note: Numbers do not sum up to 100% due to rounding and slightly differ from those provided in Table 1, as here only patients with information on all 3 mental distress indicators have been included, without missing data imputation. Abbreviations: GAD-7, Generalized Anxiety Disorder (questionnaire with 7 items); PHQ-8, Depressive symptom scale with 8 items from the Patient Health Questionnaire; SSD-12, Somatic Symptom Disorder–B Criteria Scale.

With regard to HRQoL, the sample’s mean MCS (44.5, 95%CI: 41.9 – 47.1), PCS (26.7, 95%CI: 20.2 – 33.1), and EQ-5D-5L (0.69, 95%CI: 0.60 – 0.77) scores indicated relevant (Bundesamt für Statistik (BFS), 2018; Harber-Aschan et al., 2019) impairment in mental, physical, and generic HRQoL, as compared to the general population in Switzerland, while somatic symptom burden was moderate (SSS-8: 9.5, 95%CI: 8.8 – 10.3).

### 3.2. Associations Between Sociodemographic Factors and Health Outcomes

Our analyses revealed significant associations between several sociodemographic factors and the outcomes assessed, as detailed in Tables 3 and 4. Younger age and lower income were associated with higher odds of mental distress, greater somatic symptom burden, and lower mental HRQoL (all *p-for-trend*<0.05). Younger patients showed better physical HRQoL, and those with higher income had better generic HRQoL (*p-for-trend*<0.05). Having no Swiss citizenship was associated with higher mental distress indicators, greater somatic symptom burden, and lower mental HRQoL (all *p*<0.01), while there was no indication that gender was linked to any of the outcomes. With regard to marital status, as compared to those being married or partnered, being widowed was associated with higher mental HRQoL (*p*<0.01) and less somatic symptom burden (*p*<0.05), while being divorced was associated with higher generic HRQoL (*p*<0.05). With regard to living arrangements, patients residing in a nursing home, in a hospital or other long-term care facility or other similar setting – compared to living with a spouse, partner, family or friends – exhibited lower physical (*p*<0.05) and generic HRQoL (*p*<0.01). Regarding highest educational level, lower levels were linked to lower odds of depressive distress (PHQ-8), better physical HRQoL, and better generic HRQoL (all *p-for-trend*<0.01), with associations most strongly driven by patients who did not attend school, had primary school level as highest education, or those replying with ‘not applicable’, or providing rare other replies outside primary, secondary, or tertiary levels. Finally, concerning employment, having the employment situation affected by disability, accident, physical illness, or mental illness, as compared to being employed or in training, was associated with worse outcomes throughout all 8 indicators (all *p*<0.05, with some even below lower threshold, see Tables 3 and 4). Further, being a pension recipient was linked to higher odds of exceeding the cut-off for relevant somatic distress and any of the cut-offs indicating mental distress (both *p*<0.05), being a homemaker was linked to higher odds of exceeding the cut-off for distressing anxiety symptoms (*p*<0.05), while being unemployed, looking for a job, in military service, or other rather rare employment situations was linked to higher odds of exceeding the cut-off for relevant somatic symptom distress (*p*<0.05), as compared to being employed or in training. For the sake of transparency, we provide estimates based on linear regression analyses as Online Resource 5, with estimates being largely comparable to our main analyses provided in Table 4.

**Table 3.** Results from logistic regression with the outcomes ‘scoring above cut-off’ in mental health screening instruments, predicted by sociodemographic characteristics (N = 3,179)

| Predictors | Any indicator<sup>a</sup> |  |  | PHQ<sup>b</sup> |  |  | GAD<sup>c</sup> |  |  | SSD<sup>d</sup> |  |  |
| --- | --- | --- | --- | --- | --- | --- | --- | --- | --- | --- | --- | --- |
|  | OR | lower 95%CI | upper 95%CI | OR | lower 95%CI | upper 95%CI | OR | lower 95%CI | upper 95%CI | OR | lower 95%CI | upper 95%CI |
| **Age (in years)** |  |  |  |  |  |  |  |  |  |  |  |  |
| Up to 30 | reference category |  |  | reference category |  |  | reference category |  |  | reference category |  |  |
| 30 - 39 | 0.67 | [0.28; | 1.58] | 0.78 | [0.37; | 1.61] | 0.77 | [0.36; | 1.61] | 0.77 | [0.32; | 1.80] |
| 40 - 49 | 0.70 | [0.42; | 1.15] | 0.65 | [0.38; | 1.10] | 0.85 | [0.47; | 1.50] | 0.90 | [0.59; | 1.36] |
| 50 - 59 | 0.60 | [0.31; | 1.15] | 0.59 | [0.34; | 1.02] | 0.58 | [0.23; | 1.43] | 0.68 | [0.39; | 1.18] |
| 60 - 69 | 0.46* | [0.24; | 0.87] | 0.46* | [0.26; | 0.80] | 0.41 | [0.16; | 1.03] | 0.64* | [0.41; | 0.99] |
| 70 - 79 | 0.41* | [0.21; | 0.78] | 0.45** | [0.26; | 0.77] | 0.41 | [0.16; | 1.02] | 0.56* | [0.34; | 0.90] |
| 80 and more | 0.33** | [0.19; | 0.57] | 0.39*** | [0.27; | 0.56] | 0.29* | [0.10; | 0.83] | 0.41** | [0.26; | 0.63] |
| *Test for trend: F-value (dfs) p-value* | 24.06 | (1,8) | <0.01 | 22.94 | (1,8) | <0.01 | 9.79 | (1,8) | 0.01 | 15.28 | (1,8) | <0.01 |
| **Gender** |  |  |  |  |  |  |  |  |  |  |  |  |
| Female | reference category |  |  | reference category |  |  | reference category |  |  | reference category |  |  |
| Male or other | 1.10 | [0.74; | 1.61] | 1.07 | [0.71; | 1.61] | 0.81 | [0.57; | 1.16] | 1.15 | [0.77; | 1.72] |
| **Household income (in CHF per month)** |  |  |  |  |  |  |  |  |  |  |  |  |
| Less than 4500 | reference category |  |  | reference category |  |  | reference category |  |  | reference category |  |  |
| 4500 - 5999 | 0.85 | [0.62; | 1.18] | 0.79 | [0.58; | 1.07] | 0.76 | [0.56; | 1.02] | 1.02 | [0.74; | 1.40] |
| 6000 - 8999 | 0.69* | [0.49; | 0.96] | 0.61* | [0.40; | 0.91] | 0.59* | [0.40; | 0.87] | 0.79 | [0.56; | 1.11] |
| 9000 or more | 0.56* | [0.37; | 0.84] | 0.50* | [0.29; | 0.85] | 0.48* | [0.29; | 0.80] | 0.69 | [0.44; | 1.07] |
| *Test for trend: F-value (dfs) p-value* | 18.22 | (1,8) | <0.01 | 12.35 | (1,8) | <0.01 | 15.50 | (1,8) | <0.01 | 7.27 | (1,8) | 0.03 |
| **Nationality** |  |  |  |  |  |  |  |  |  |  |  |  |
| Swiss citizenship | reference category |  |  | reference category |  |  | reference category |  |  | reference category |  |  |
| No Swiss citizenship | 1.33*** | [1.20; | 1.48] | 1.41*** | [1.26; | 1.57] | 1.59** | [1.23; | 2.06] | 1.33** | [1.11; | 1.60] |
| **Marital status** |  |  |  |  |  |  |  |  |  |  |  |  |
| Divorced or dissolved partnership | 1.07 | [0.80; | 1.42] | 1.13 | [0.87; | 1.47] | 1.20 | [0.70; | 2.03] | 0.94 | [0.64; | 1.37] |
| Married or partnered | reference category |  |  | reference category |  |  | reference category |  |  | reference category |  |  |
| Single or unmarried | 1.07 | [0.82; | 1.40] | 1.03 | [0.79; | 1.33] | 1.09 | [0.74; | 1.61] | 1.01 | [0.62; | 1.64] |
| Widowed | 0.89 | [0.57; | 1.38] | 0.80 | [0.48; | 1.32] | 0.93 | [0.57; | 1.51] | 0.79 | [0.48; | 1.31] |

| Living arrangement |  |  |  |  |  |  |  |  |  |  |  |  |
| --- | --- | --- | --- | --- | --- | --- | --- | --- | --- | --- | --- | --- |
| With spouse/partner/family/friends | reference category |  |  | reference category |  |  | reference category |  |  | reference category |  |  |
| Alone | 1.00 | [0.79; 1.28] |  | 1.10 | [0.87; 1.39] |  | 1.01 | [0.65; 1.56] |  | 1.09 | [0.78; 1.51] |  |
| In a nursing home / hospital or other long-term care facility / other | 1.45 | [0.69; 3.03] |  | 0.99 | [0.41; 2.37] |  | 1.20 | [0.41; 3.50] |  | 1.53 | [0.79; 2.95] |  |
| Highest educational level |  |  |  |  |  |  |  |  |  |  |  |  |
| Did not attend school / primary level / not applicable / other | 0.62 | [0.36; 1.04] |  | 0.52* | [0.31; 0.86] |  | 0.74 | [0.37; 1.44] |  | 0.94 | [0.57; 1.53] |  |
| Lower secondary | 0.85 | [0.55; 1.30] |  | 0.86 | [0.61; 1.23] |  | 0.89 | [0.56; 1.42] |  | 0.98 | [0.64; 1.50] |  |
| Upper secondary | 0.89 | [0.70; 1.13] |  | 0.89 | [0.68; 1.15] |  | 0.95 | [0.70; 1.29] |  | 1.00 | [0.82; 1.22] |  |
| Tertiary level | reference category |  |  | reference category |  |  | reference category |  |  | reference category |  |  |
| Test for trend: F-value (dfs) p-value | 4.77 | (1,8) | 0.06 | 12.67 | (1,8) | <0.01 | 1.36 | (1,8) | 0.28 | 0.09 | (1,8) | 0.77 |
| Employment situation |  |  |  |  |  |  |  |  |  |  |  |  |
| Employed or in training | reference category |  |  | reference category |  |  | reference category |  |  | reference category |  |  |
| Pension recipient | 1.49* | [1.07; 2.06] |  | 1.20 | [0.83; 1.72] |  | 1.22 | [0.77; 1.92] |  | 1.70* | [1.17; 2.48] |  |
| Homemaker | 1.64 | [0.91; 2.97] |  | 1.50 | [0.87; 2.57] |  | 1.93* | [1.07; 3.47] |  | 1.93 | [0.94; 3.97] |  |
| Disability / accident / physical illness / mental illness | 2.76*** | [1.97; 3.88] |  | 2.32** | [1.64; 3.27] |  | 2.31** | [1.54; 3.48] |  | 3.31*** | [2.35; 4.68] |  |
| Unemployed / looking for a job / in military service / other | 1.36 | [0.87; 2.11] |  | 1.12 | [0.60; 2.09] |  | 1.42 | [0.82; 2.45] |  | 1.87* | [1.03; 3.37] |  |

<sup>a</sup>50 missing values; <sup>b</sup>20 missing values; <sup>c</sup>36 missing values; <sup>d</sup>29 missing values.
\* $p < 0.05$ ; \*\* $p < 0.01$ ; \*\*\* $p < 0.001$ .
Notes: Analyses conducted taking into account the sampling design and applying multiple imputation to handle missing data.
Abbreviations: CHF, Swiss Francs; CI, confidence interval; GAD, Generalized Anxiety Disorder (questionnaire with 7 items); PHQ, Depressive symptom scale with 8 items from the Patient Health Questionnaire; SSD, Somatic Symptom Disorder–B Criteria Scale.

**Table 4.** Results from Generalized Linear Model regression analyses, predicting the continuous variables MCS, PCS, SSS-8, and EQ-5D-5L by sociodemographic characteristics (N = 3,179)

| Predictors | Scaled MCS<sup>a</sup> |  |  | Scaled PCS<sup>b</sup> |  |  | SSS-8<sup>c</sup> |  |  | Transformed EQ-5D-5L<sup>d</sup> |  |  |
| --- | --- | --- | --- | --- | --- | --- | --- | --- | --- | --- | --- | --- |
|  | Coefficients | lower 95%CI | upper 95%CI | Coefficients | lower 95%CI | upper 95%CI | Coefficients | lower 95%CI | upper 95%CI | Coefficients | lower 95%CI | upper 95%CI |
| **Age (in years)** |  |  |  |  |  |  |  |  |  |  |  |  |
| Up to 30 | reference category |  |  | reference category |  |  | reference category |  |  | reference category |  |  |
| 30 - 39 | 0.05 | [-0.04; 0.14] |  | 0.00 | [-0.07; 0.06] |  | -0.11 | [-0.27; 0.05] |  | 0.00 | [-0.07; 0.08] |  |
| 40 - 49 | 0.05 | [-0.03; 0.13] |  | -0.02 | [-0.13; 0.09] |  | -0.11* | [-0.22; 0] |  | 0.06 | [-0.01; 0.13] |  |
| 50 - 59 | 0.09 | [-0.01; 0.18] |  | -0.11 | [-0.27; 0.05] |  | -0.16* | [-0.31; -0.01] |  | 0.05 | [-0.03; 0.12] |  |
| 60 - 69 | 0.13* | [0.01; 0.24] |  | -0.16 | [-0.40; 0.08] |  | -0.26** | [-0.45; -0.08] |  | 0.03 | [-0.05; 0.12] |  |
| 70 - 79 | 0.14* | [0.02; 0.27] |  | -0.24 | [-0.50; 0.01] |  | -0.23** | [-0.39; -0.08] |  | 0.01 | [-0.07; 0.10] |  |
| 80 and more | 0.15* | [0.02; 0.28] |  | -0.32* | [-0.55; -0.09] |  | -0.25** | [-0.37; -0.12] |  | 0.01 | [-0.07; 0.08] |  |
| *Test for trend: F-value (dfs) p-value* | 8.82 | (1,8) | 0.02 | 8.62 | (1,8) | 0.02 | 23.27 | (1,8) | <0.01 | 0.00 | (1,8) | 0.96 |
| **Gender** |  |  |  |  |  |  |  |  |  |  |  |  |
| Female | reference category |  |  | reference category |  |  | reference category |  |  | reference category |  |  |
| Male or other | 0.00 | [-0.06; 0.05] |  | 0.01 | [-0.14; 0.15] |  | -0.03 | [-0.18; 0.12] |  | 0.05 | [-0.01; 0.11] |  |
| **Household income (in CHF per month)** |  |  |  |  |  |  |  |  |  |  |  |  |
| Less than 4500 | reference category |  |  | reference category |  |  | reference category |  |  | reference category |  |  |
| 4500 - 5999 | 0.03 | [-0.02; 0.08] |  | 0.03 | [-0.03; 0.10] |  | -0.06 | [-0.15; 0.04] |  | 0.03 | [-0.01; 0.07] |  |
| 6000 - 8999 | 0.05 | [-0.01; 0.10] |  | 0.06 | [-0.01; 0.14] |  | -0.07 | [-0.17; 0.03] |  | 0.03* | [0; 0.07] |  |
| 9000 or more | 0.08* | [0.01; 0.15] |  | 0.06 | [-0.02; 0.14] |  | -0.15* | [-0.27; -0.02] |  | 0.05** | [0.02; 0.09] |  |
| *Test for trend: F-value (dfs) p-value* | 10.55 | (1,8) | 0.01 | 4.00 | (1,8) | 0.08 | 10.16 | (1,8) | 0.01 | 16.52 | (1,8) | <0.01 |
| **Nationality** |  |  |  |  |  |  |  |  |  |  |  |  |
| Swiss citizenship | reference category |  |  | reference category |  |  | reference category |  |  | reference category |  |  |
| No Swiss citizenship | -0.04** | [-0.07; -0.01] |  | -0.03 | [-0.08; 0.02] |  | 0.10* | [0.03; 0.17] |  | -0.02 | [-0.05; 0.02] |  |
| **Marital status** |  |  |  |  |  |  |  |  |  |  |  |  |
| Divorced or dissolved partnership | -0.01 | [-0.05; 0.04] |  | 0.00 | [-0.09; 0.09] |  | 0.05 | [-0.03; 0.13] |  | 0.03* | [0; 0.06] |  |
| Married or partnered | reference category |  |  | reference category |  |  | reference category |  |  | reference category |  |  |
| Single or unmarried | 0.00 | [-0.04; 0.04] |  | 0.00 | [-0.09; 0.09] |  | -0.01 | [-0.14; 0.12] |  | 0.01 | [-0.05; 0.07] |  |
| Widowed | 0.07** | [0.02; 0.11] |  | -0.03 | [-0.12; 0.06] |  | -0.10* | [-0.18; -0.01] |  | 0.04 | [-0.01; 0.09] |  |

| Living arrangement |  |  |  |  |  |  |  |  |  |  |  |  |
| --- | --- | --- | --- | --- | --- | --- | --- | --- | --- | --- | --- | --- |
| With spouse/partner/family/friends | reference category |  |  | reference category |  |  | reference category |  |  | reference category |  |  |
| Alone | -0.01 | [-0.05; | 0.02] | -0.02 | [-0.09; | 0.05] | 0.01 | [-0.09; | 0.10] | -0.03 | [-0.06; | 0.01] |
| In a nursing home / hospital or other long-term care facility / other | -0.06 | [-0.19; | 0.07] | -0.20* | [-0.39; | -0.02] | 0.08 | [-0.12; | 0.27] | -0.18** | [-0.31; | -0.06] |
| Highest educational level |  |  |  |  |  |  |  |  |  |  |  |  |
| Did not attend school / primary level / not applicable / other | 0.02 | [-0.02; | 0.06] | 0.14* | [0.04; | 0.25] | -0.04 | [-0.18; | 0.10] | 0.08* | [0.01; | 0.15] |
| Lower secondary | 0.01 | [-0.07; | 0.08] | 0.07 | [-0.01; | 0.16] | 0.03 | [-0.07; | 0.13] | 0.07*** | [0.04; | 0.10] |
| Upper secondary | 0.01 | [-0.02; | 0.05] | 0.02 | [-0.05; | 0.08] | -0.01 | [-0.07; | 0.06] | 0.04** | [0.02; | 0.07] |
| Tertiary level | reference category |  |  | reference category |  |  | reference category |  |  | reference category |  |  |
| Test for trend: F-value (dfs) p-value | 1.56 | (1,8) | 0.25 | 13.80 | (1,8) | <0.01 | 0.19 | (1,8) | 0.67 | 12.60 | (1,8) | <0.01 |
| Employment situation |  |  |  |  |  |  |  |  |  |  |  |  |
| Employed or in training | reference category |  |  | reference category |  |  | reference category |  |  | reference category |  |  |
| Pension recipient | -0.03 | [-0.09; | 0.03] | -0.05 | [-0.12; | 0.03] | 0.06 | [-0.05; | 0.17] | 0.00 | [-0.04; | 0.04] |
| Homemaker | -0.07 | [-0.15; | 0.01] | -0.05 | [-0.19; | 0.10] | 0.12 | [-0.04; | 0.28] | -0.03 | [-0.08; | 0.02] |
| Disability / accident / physical illness / mental illness | -0.15** | [-0.22; | -0.08] | -0.25*** | [-0.36; | -0.15] | 0.23** | [0.13; | 0.33] | -0.12* | [-0.21; | -0.03] |
| Unemployed / looking for a job / in military service / other | -0.04 | [-0.12; | 0.03] | -0.06 | [-0.17; | 0.04] | -0.01 | [-0.18; | 0.16] | -0.04 | [-0.09; | 0.02] |

<sup>a</sup>Scaled *T*-scores – 50 missing values – Gaussian GLMs with log link function; <sup>b</sup>Scaled *T*-scores – 30 missing values – Gaussian GLMs with log link function; <sup>c</sup>Raw score – 36 missing values – Gamma distribution GLMs with log link function; <sup>d</sup>Scaled score – 29 missing values – Binomial distribution GLMs with probit link function.
\* $p < 0.05$ ; \*\* $p < 0.01$ ; \*\*\* $p < 0.001$ .
Notes: Analyses conducted taking into account the sampling design and applying multiple imputation to handle missing data.
Abbreviations: CHF, Swiss Francs; CI, confidence interval; EQ-5D, European Quality of Life-5 Dimensions questionnaire; MCS, Mental Component Summary of the SF-36; PCS, Physical component summary of the SF-36; SSS-8, Somatic Symptom Scale-8.

## 4. Discussion

### 4.1. Main findings

In this large multicenter sample of somatic hospital inpatients, over one-third of participants met cut-off criteria for at least one domain of mental distress (depressive, anxiety, or somatic symptom distress). Notably, 19% of patients from our sample fulfilled multiple criteria, reflecting a considerable burden of multimodal mental distress. Our results also highlight significant impairments in mental, physical, and generic HRQoL, compared to general population norms. Although our participants’ average somatic symptom burden was in a moderate range, it still exceeded community benchmarks from previous research.

Regarding sociodemographic characteristics, younger age, lower income, and non-Swiss citizenship each independently was associated with higher odds of mental distress, greater somatic symptom burden, and/or worse mental HRQoL. Younger patients, however, demonstrated better physical HRQoL. Employment situations marked by disability, accident, or chronic illness likewise were associated with higher likelihood of meeting mental-health cut-offs, alongside lowered mental, physical, and generic HRQoL. Notably, there was a somewhat unexpected pattern, indicating that regarding lower highes education levels were linked to lower odds of depressive symptoms, to better physical HRQoL, and to better generic HRQoL. Other factors –gender, marital status, and living arrangement – showed no or inconsistent patterns, except for specific subgroups.

### 4.2. Comparison with previous studies

Our prevalence estimates and distribution of mental distress align broadly with earlier findings, yet appear higher in some domains. For example, a meta-analysis of general medical and surgical inpatients estimated that 5–34% of patients showed clinically relevant depressive symptoms, with an average of 12% [45], while others reported pooled anxiety-symptom prevalence of 28% and anxiety disorder of 8% in general hospital inpatients [46]. Our combined depression-anxiety estimates place us at the upper end of these ranges, comparable to other studies suggesting high levels of psychosocial distress among medically ill cohorts [47] and corroborating evidence from non-psychiatric hospital departments, identifying 34% of patients as cases with mental disorders [48]. Somatic symptom distress is less commonly assessed in large inpatient samples; however, our rate of 25% parallels an exploratory finding of about 22% for “bodily distress disorder” [49].

Our study identified associations between various sociodemographic factors and the prevalence of mental distress, a finding that complements existing literature. The lower risk of mental distress with advancing age parallels findings from other inpatient samples, where older age has been linked to reduced odds of acute mental-somatic multimorbidities [50, 51]. This may reflect more adaptive coping mechanisms, selective survival, or differential detection of distress among older adults. In contrast, younger adults may face more mental challenges during hospitalization, consistent with studies showing increased anxiety in younger hospitalized cohorts [51]. Our observation that lower income was independently associated with worse mental-health outcomes confirms extensive literature linking financial strain to heightened psychosocial vulnerability, including in inpatients [52–54].

Patients without Swiss citizenship manifested higher odds of all investigated forms of mental and somatic distress and lower mental HRQoL. This aligns with reports of increased psychosocial and mental distress among migrants, potentially driven by language barriers, reduced social support, discrimination, and legal uncertainties [55–57]. The category “patients without Swiss citizenship” encompasses a heterogeneous spectrum of individuals, ranging from nationalities often representing educated expatriates typically employed in professional or academic contexts to economically vulnerable labor migrants with lower-income positions conditions. Future studies should consider stratifying this group further to inform culturally and socioeconomically tailored interventions to improve health outcomes within these subpopulations.

Contrary to many community-based studies highlighting a female predominance in depression, anxiety, and somatic symptom burden [58–60] our data did not reveal a gender association with mental distress or HRQoL. Similar null findings have been previously reported [50, 61] suggesting that gender differences may be overshadowed by the severity of medical conditions in acute settings.

Interestingly, we found that being widowed was associated with better mental HRQoL and less somatic symptom burden – a result potentially reflecting ongoing bereavement adjustments, less burden by spouse caregiving, especially in older participants, or a selection effect of individuals with higher resilience [62]. At the same time, being divorced or in dissolved partnership was associated with improved generic HRQoL, again, potentially hinting at unmeasured factors – such as social support or coping strategies – specific to that subgroup.

The observed association of residing in nursing homes/ long-term facilities with lower physical and generic HRQoL aligns with established evidence of heightened frailty, multimorbidity, and functional decline among these individuals [63]. However, our data did not suggest that nursing home residents were systematically more prone to anxiety or depressive symptoms, highlighting the potentially complex interplay among social supports, institutional resources, and physical care needs. Of note, 95%CIs of these predictions were rather large, hence respective estimates were rather imprecise.

A particularly unexpected finding was that patients reporting lower or minimal formal education had lower odds of relevant depressive symptoms and somewhat better physical and generic HRQoL scores than those with tertiary-level education. This pattern contrasts with most epidemiological research [64–66]. However, results from previous studies indicate that this association appears to decrease with age [67] and in some cases lowest mental HRQoL was not reported in low but in middle educated subjects [68]. Notably, our findings may also indicate higher resilience towards distress related to hospitalization in patients with lower education. Alternatively, individuals with higher education may be more prone to self-reporting subtle psychological symptoms or may bring more complex healthcare expectations. Future work is needed to clarify how educational attainment modifies resilience and reporting behaviors in somatic inpatient care settings.

Consistent with prior research [69], disability-related employment situations were associated with higher mental distress odds across virtually all indicators. The cumulative burden of chronic illness, social stigma, and potential economic insecurity likely underlies these associations. Moreover, pension recipients also demonstrated elevated odds of ‘any mental health distress’ and being above somatic distress cut-offs. By contrast, previous evidence indicated that statutory and voluntary retirement is linked to higher mental health, although the improvement seems to attenuate over time [70]. However, opposite findings were reported following retirement due to ill health [70], indicating that being on early or partial pension could imply vulnerability in somatic inpatients. By contrast, ‘homemaker’ status was specifically linked to anxiety, while other less common employment categories were linked to somatic distress – highlighting the complex interplay between occupational roles, psychosocial stressors, and physical conditions in hospitalized populations.

Taken together, our findings corroborate the multidimensional nature of mental distress in somatic hospital settings while extending current knowledge about how specific demographic and socioeconomic subgroups may be at heightened risk. They reinforce the importance of proactive screening, integrated psychosomatic or psychiatric care, and, ultimately, tailored interventions that account for each patient’s age, cultural and linguistic background, socioeconomic status, and employment circumstances.

### 4.3. Strengths and Limitations

Our study has several strengths. First, we recruited a large and diverse sample of hospital inpatients, thereby enhancing the generalizability and robustness of our findings. Second, our use of a multidimensional mental assessment is a key strength. By incorporating a wide range of psychological measures, we obtained a thorough and nuanced overview of the mental health challenges faced by somatic hospital inpatients, ensuring that multiple aspects of mental distress were accurately captured. Third, stratifying data by demographic variables offers deeper insights into how mental health issues differ across subgroups, aiding in the identification of high-risk populations and informing targeted interventions.

Our study also has several limitations: First, we did not include the full range of medical specialties. For example we did not include surgical wards (some patients, e.g., from gynecology wards may still have undergone surgery) or wards with patients primarily treated for oncological conditions, as for these patients, specific psychosocial support services are already available. This limits the generalizability of our findings to related hospital inpatient populations, as for example surgical inpatients might exhibit different patterns or levels of mental distress compared to other somatic inpatients. Additionally, the inclusion of gynecology patients may have introduced a gender bias in our sample, potentially affecting the study findings, even though we adjusted all analyses for gender. Moreover, a putative selection bias toward fitter patients, particularly within the geriatric population, could have affected the results, as these patients may report lower levels of distress and higher HRQoL compared to those who are less fit. Our study methods imply potential sources of imprecision, for instance, the reliance on self-assessment questionnaires, since it was not feasible to conduct standardized clinical interviews due to the large number of patients and resource constraints. Although comprehensive, this may be subject to response biases, such as social desirability or recall bias.

### 4.4. Generalizability

The generalizability of our study findings, conducted in a city within Switzerland’s highly developed medical system, is an important aspect to consider. The advanced healthcare infrastructure and high standard of medical care in Switzerland may influence the prevalence and management of mental distress and HRQoL in somatic hospital inpatients, potentially differing from settings with less developed healthcare systems. Additionally, the study’s urban setting and inclusion of tertiary somatic hospitals could also affect its external validity, as respective healthcare facilities often have more specialized services and resources, which might not be readily available in rural or less populated areas. This disparity could lead to differences in patient outcomes, experiences, and the prevalence of mental distress. Cultural factors inherent to the Swiss population, such as attitudes towards mental health and disease, help-seeking behaviors, and stigma, may not be representative of populations in other countries. These cultural characteristics may play a relevant role in the manifestation and reporting of mental distress, potentially influencing the study findings.

We cannot exclude bias due to selective participation. Registry-based comparisons revealed a modest selection bias: our cohort contained about four percentage-points more men and proportionally fewer patients aged ≥ 70 years than the source population. These departures likely reflect shorter average stays on the gynecology wards and the practical difficulty of recruiting frailer older adults. While the differences are small, they may slightly overestimate mental-health prevalence in younger males and underestimate it in older females, limiting strict generalizability. Future studies should incorporate targeted approaches – especially on short-stay and geriatric wards – to strengthen representativeness and minimize such bias.

Taken together, while our study provides valuable insights into the prevalence and characteristics of mental distress, somatic symptoms, and HRQoL among somatic inpatients in a Swiss urban context, caution should be exercised in extrapolating these findings to other settings. Further studies, including surgical inpatients, in diverse geographical and healthcare settings are needed to enhance the generalizability of our findings.

### 4.5. Clinical, public health, and future research implications

The clinical and public health implications of our findings are considerable. From a health-services perspective, the identified social gradients point out to actionable levers. They highlight the need for heightened awareness and systematic strategies to detect and address mental distress among somatic inpatients – particularly those at elevated risk due to specific sociodemographic factors [71, 72]. Proactive integrated psychosocial CL models have emerged as a promising approach in this context [73], showing benefits for older patients, clinical teams, and hospital systems, as demonstrated in the HOME study [21]. Our results support the incorporation of routine mental distress screening into standard inpatient care to facilitate early identification and timely intervention, with the potential to improve both individual outcomes and system-level efficiency.

In terms of public health, our study highlights the importance of integrating such proactive integrated mental health CL services into general healthcare, especially in hospital settings, to address the comprehensive needs of patients. For example, guided internet-based acceptance and commitment therapy (ACT) improved somatic, depressive, and anxiety symptoms in adults with persistent physical symptoms in a randomised trial [74], suggesting scalable, low-threshold adjuncts to stepped care.

From a research perspective, our study underscores the necessity for further investigations in diverse healthcare settings and populations, including surgical wards and regions with different healthcare infrastructures and cultural backgrounds. Future research should aim to establish more definitive causal relationships of sociodemographic factors with mental distress, somatic symptoms, and HRQoL, and explore the effectiveness of various intervention strategies. It is also crucial to investigate the long-term impact of mental distress on patient outcomes and healthcare utilization.

### 4.6 Conclusion

Our findings underscore the high frequency of mental distress among inpatients in tertiary somatic hospitals and reveal that one in three exhibits clinically relevant depression, anxiety, or somatic symptom distress – often with overlap. Mental, physical, and generic HRQoL were notably reduced, emphasizing the extensive burden of mental-somatic multimorbidity in this setting. Younger age, lower socioeconomic status, and non-Swiss citizenship emerged as important correlates of increased risk, indicating that psychosocial screening and interventions should be carefully tailored to at-risk subgroups. These results highlight the urgent need for proactive psychosomatic and psychiatric CL services – such as SCCMs – that seamlessly integrate mental healthcare into routine hospital treatment. By addressing the complex interplay of mental and physical health, such models can better meet patients’ comprehensive needs, potentially improving health outcomes, patient satisfaction, and healthcare efficiency. Ongoing analyses, based on data from the SomPsyNet study will assess the impact and cost-effectiveness of the SCCM, expected to offer valuable insights into optimizing mental healthcare provision in somatic hospital settings.

## Supporting information

Online Resource 1 - Sample Comparison

Online Resource 2 - Sociodemographics

Online Resource 3 - Supplemental Methods

Online Resource 4 - Figure Flow Chart

Online Resource 5 - Results from Linear Regression

## Data Availability

The datasets being held by the SomPsyNet project are not readily available due to their sensitive nature. In the case of inquiries by third parties that wish to reuse data SomPsyNet data after an embargo period, the following procedure is planned. Researchers interested in the data may submit a project synopsis addressed to the publications committee of the SomPsyNet project and will have to obtain authorization from the responsible ethics committee as ordained in the Ordinance of 20 September 2013 on Human Research with the exception of Clinical Trials (The Swiss Federal Council, 2013) (Human Research Ordinance, HRO). The publication committee will review the project synopsis and will answer the formal requests of applicants. Only upon collection of all important consents and upon approval of the responsible ethics committee(s), the requested data will be transferred to the applicants. Third parties must confirm and provide evidence to comply with all relevant Swiss and cantonal laws and regulations (especially regarding data protection and Human Research), as well as all obligations and regulations set out in the documents and contracts related to SomPsyNet. Fees may apply to cover expenses related to data reuse. Requests to access the datasets should be directed to Gunther Meinlschmidt. Any future changes to the data sharing plan will be noted in Data Availability Statements and updated in the registry record.

## Members of the SomPsyNet Consortium

Members of the SomPsyNet consortium: Bachmann, M., Department of Psychosomatic Medicine and Psychotherapy, Klinik Barmelweid AG, Barmelweid, Switzerland; Baenteli, I., Department of Psychosomatic Medicine, University Hospital and University of Basel, Basel, Switzerland; Bahmane, S., Department of Psychosomatic Medicine, University Hospital and University of Basel, Basel, Switzerland; Bales, G., University Department of Geriatric Medicine FELIX PLATTER, Basel, Switzerland; Bally, K., Centre for Primary Health Care, University of Basel, Switzerland; Bassetti, S., Division of Internal Medicine, University Hospital and University of Basel, Basel, Switzerland, Department of Clinical Research, University Hospital and University of Basel, Basel, Switzerland; Baumgartner, R., Social Insurance Institution Basel-Landschaft, Binningen, Switzerland; Beck, J., Clinic Sonnenhalde AG, Riehen, Switzerland; Bosman, S., Department of Psychosomatics and Psychiatry, Bethesda Hospital, Basel, Switzerland; Buechel, D., University of Basel c/o University Hospital of Basel, Department of Clinical Research, Basel, Basel-Stadt, Schweiz; Dörner, A., St. Claraspital, Medical clinic, Basel, Switzerland; Ebner, L., Department of Psychosomatic Medicine, University Hospital and University of Basel, Basel, Switzerland; Erb, J., Department of Psychosomatic Medicine, University Hospital and University of Basel, Basel, Switzerland; Fink, G., Swiss Tropical and Public Health Institute, Basel, Switzerland; University of Basel, Basel, Switzerland; Frick, A., Department of Psychosomatic Medicine, University Hospital and University of Basel, Basel, Switzerland; Fuchs, S., Department of Health Canton Basel-Stadt, Medical Services, Basel, Switzerland; Grossmann, F., Department of Medicine, Division of Nursing, University Hospital Basel, Basel, Switzerland; Hermann, A., Direktion Pflege/MTT, University Hospital and University of Basel, Basel, Switzerland; Hotopf, M., Department of Psychological Medicine, Institute of Psychiatry, Psychology and Neuroscience, King’s College London, London, United Kingdom; Huber, C., University Psychiatric Clinics (UPK), University of Basel, Basel, Switzerland; Isler-Christ, L., Sevogel-Apotheke, Basel, Switzerland, Baselstädtischer Apotheker-Verband, Basel, Switzerland; Karpf, C., Department of Health Canton Basel-Stadt, Division of Prevention, Basel, Switzerland; Katapodi, MC., Department of Clinical Research, University of Basel, Basel, Switzerland, University of Michigan School of Nursing, Ann Arbor, MI USA; Keller, RC., Swiss Heart Foundation, Bern, Switzerland; Klimmeck, S., University Hospital Basel, Basel, Switzerland; Lang, UE., University Psychiatric Clinics (UPK), Department of Psychiatry and Psychotherapy, Basel, Switzerland; Mazander, S., IV-Stelle Basel-Stadt, Basel, Switzerland; Meinlschmidt, G., Department of Digital and Blended Psychosomatics and Psychotherapy, Psychosomatic Medicine, University Hospital and University of Basel, Basel, Switzerland, Department of Psychosomatic Medicine, University Hospital and University of Basel, Basel, Switzerland, Trier University, Department of Clinical Psychology and Psychotherapy – Methods and Approaches, Trier, Germany; Schaefert, R., Department of Psychosomatic Medicine, University Hospital and University of Basel, Basel, Switzerland; Schirmer, F., Vereinigung der psychosomatisch tätigen Aerztinnen und Aerzte der Region Basel, Basel, Switzerland; Schur, N., Institute of Pharmaceutical Medicine (ECPM), University of Basel, Basel, Switzerland; Schwenkglenks, M., Institute of Pharmaceutical Medicine (ECPM), University of Basel, Basel, Switzerland; Health Economics Facility, Department of Public Health, University of Basel, Basel, Switzerland; Schwob, P., Psychotherapists Association of Basel VPB, Basel, Switzerland; Seelmann, SF., Department of Internal Medicine, University Hospital Basel, Basel, Switzerland; Stiefel, F., Liaisonpsychiatrischer Dienst, University Hospital Lausanne, Lausanne, Switzerland; Studer, A., Department of Health Canton Basel-Stadt, Division of Prevention, Basel, Switzerland; Tegethoff, M., Institute of Psychology, RWTH Aachen University, Aachen, Germany; Trost, S., Department of Geriatric Psychiatry, Universitäre Altersmedizin FELIX PLATTER, Basel, Switzerland; Tschudin, S., Department of Obstetrics and Gynecology, University Hospital and University of Basel, Switzerland; Urech, C., Gyn. Social Medicine and Psychosomatics, University Hospital and University of Basel, Basel, Switzerland; Weyermann, D., Patientenstelle Basel, Basel, Switzerland; Wyss, K., Swiss Tropical and Public Health Institute, Basel, Switzerland; University of Basel, Basel, Switzerland; Zwahlen, D., Department of Psychosomatic Medicine, University Hospital and University of Basel, Basel, Switzerland; von Allmen, T., Department of Health Canton Basel-Stadt, Health Care, Basel, Switzerland.

## Acknowledgments

The authors thank all patients who participated in the study and generously shared their experiences, as well as all members of the consortium involved in dedicating support in patient recruitment at the various study sites. We are grateful to all master students involved for their assistance in recruitment and overall study implementation. Special thanks go to the patient participation committee for their valuable input in grant preparation, study design, refinement of consent materials, and strategies for enrolment and dissemination. We thank the physicians, nursing and other staff from all participating units at each study site for their support throughout the study. We also acknowledge the contributions of the involved ICT specialists, whose technical support was essential.

## Author contributions

The coordinating center at UHB had the role of overseeing all activities at all sites. The steering committee of SomPsyNet consisted of the project head and responsible of operations of the Department of Health Canton Basel-Stadt, Division of Prevention, the study sponsor, principal investigators, and project responsible of operations at the coordinating center, as well as a representative of St. Claraspital Medical Clinic (CLARA) that took part in the SomPsyNet project but not the SomPsyNet study. The steering committee had the role of deciding upon all major aspects of the study. The endpoint was discussed among the investigators of the steering committee, with input from other members of the co-author group, with some of them (K.W., G.F., M.S.) being members of the external evaluation board, evaluating SomPsyNet and being implemented by Gesundheitsförderung Schweiz (GFCH), the main funding body of SomPsyNet. An advisory board provided guidance and feedback related to the project and the study. A patient advisory group consisting of several patient representatives worked together to oversee and give feedback on study material and protocol. The SomPsyNet consortium met at least on a yearly basis, discussing the development of the study, and giving critical feedback on the study conduction and necessary adaptations.

C.K., R.S., A.F., and GM first identified the question leading to the formation of this research. G.M., A.F., C.K., A.S., I.B., M.B., A.D., S.Ts., K.W., G.F., M.S., R.S. contributed to the development of the main trial protocol. RS contributed significantly also as the sponsor-investigator of this study. G.M. served as lead principal investigator and A.F., M.B., R.S., S.Tr., and S.Ts. served as principal investigators at the different study sites, contributing significantly to data collection and protocol adherence. I.B. and A.S. acted as study operative leads, overseeing and managing the execution of the study. G.M. drafted the first version of the manuscript. S.B. and V.O. conducted statistical analyses. I.B., L.E., and D.B. were in charge and conducted data management tasks. All authors made significant revisions to the manuscript for important intellectual content and all authors reviewed and approved the final version of the manuscript for submission, reflecting their agreement with the work in its current form and their acceptance of accountability for all aspects of the work. This manuscript was written following the STROBE reporting guidelines (Von Elm et al., 2007). We used artificial intelligence (AI)-based tools, including ChatGPT and OpenEvidence to support manuscript preparation. We used publicly available search technologies, which we recognize likely utilise AI capabilities. We confirm that the contributions of AI were strictly in an assistive capacity. AI was not involved in conceptual tasks. Human oversight was continuously employed to ensure the accuracy of content and address any ethical concerns.

## Ethical standards

All procedures complied with institutional/national standards, the Helsinki declaration of 1975, as revised in 2008, and the Human Research Act. Written informed consent was obtained. The *Ethikkommission Nordwest- und Zentralschweiz* (EKNZ; No. 2019–01724) approved the study (and four amendments). The study is registered with the Swiss National Clinical Trials Portal and ClinicalTrials.gov (NCT04269005).

## Captions of supplementary online information

*Online Resource 1* – eTable: Comparison of the SomPsyNet Study Population with Hospital Registry Data, Regarding Patients’ Gender and Age

*Online Resource 2* – eTable: Sociodemographic characteristics of the study sample

*Online Resource 3* – Supplemental Methods

*Online Resource 4* – eFigure: Flowchart of the study sample

*Online Resource 5* – eTable: Results from linear regression analyses, predicting the continuous variables MCS, PCS, SSS-8, and EQ-5D-5L by sociodemographic characteristics (N = 3,179)

STROBE Statement—checklist of items that should be included in reports of observational studies

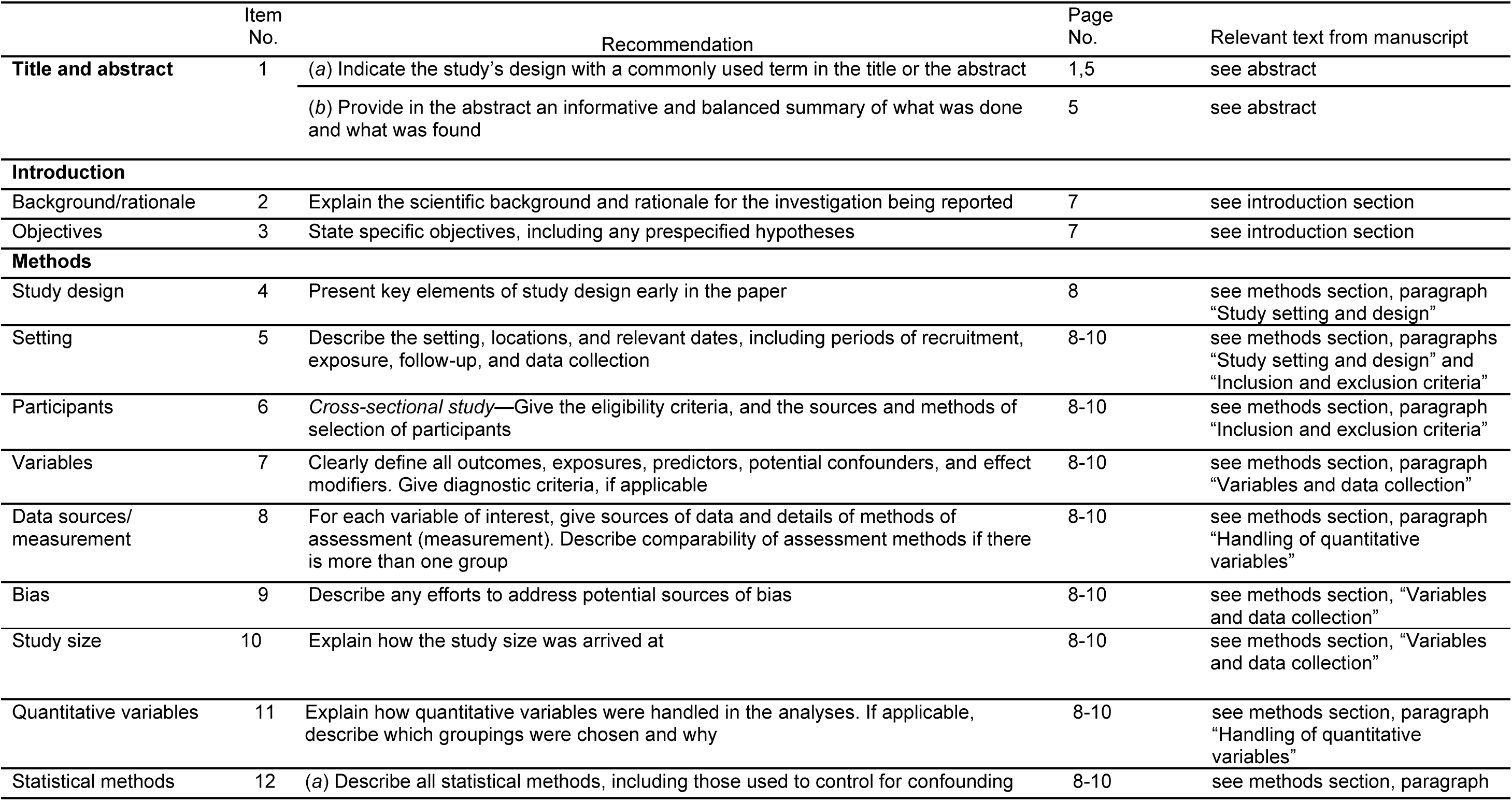
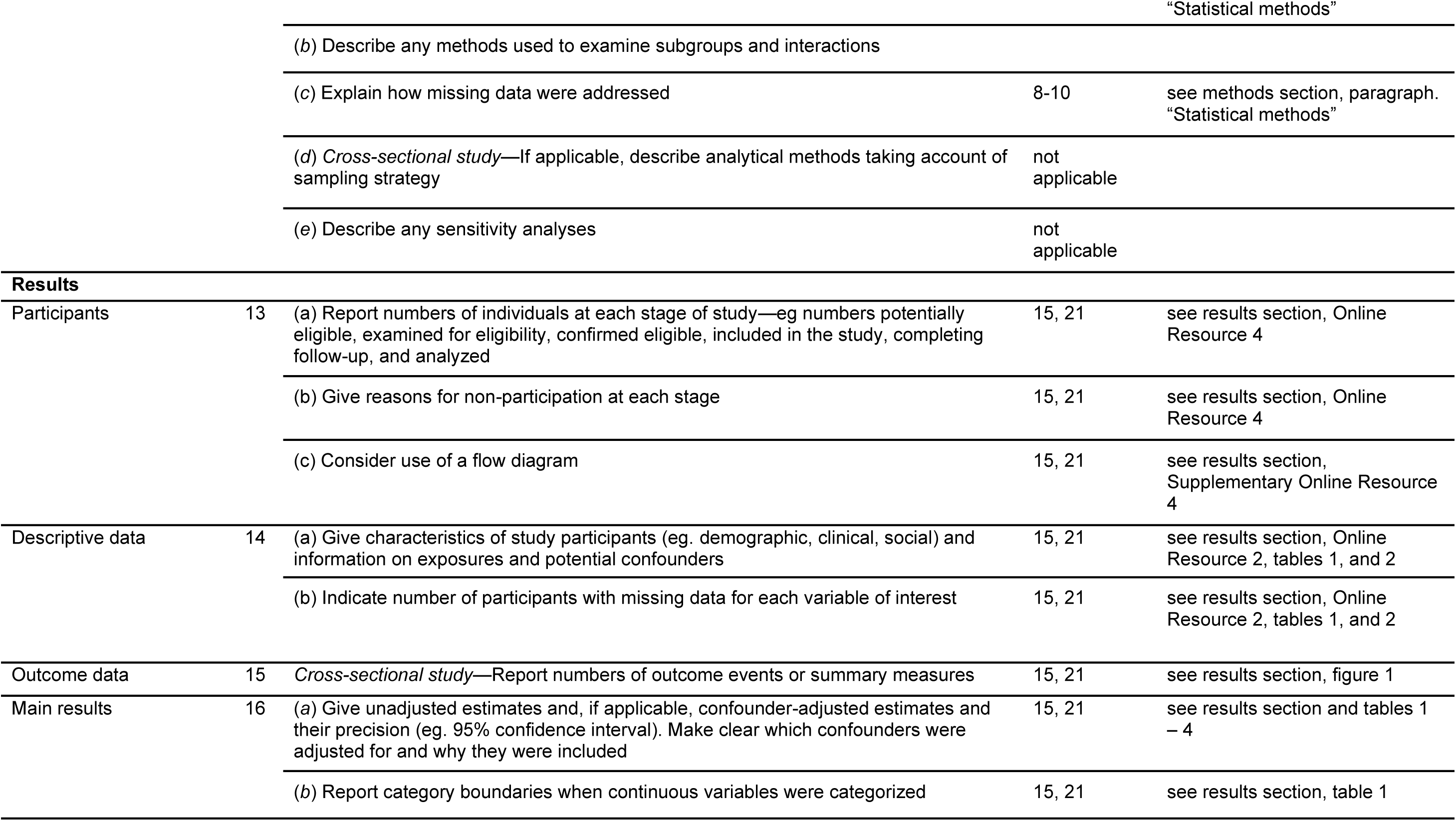
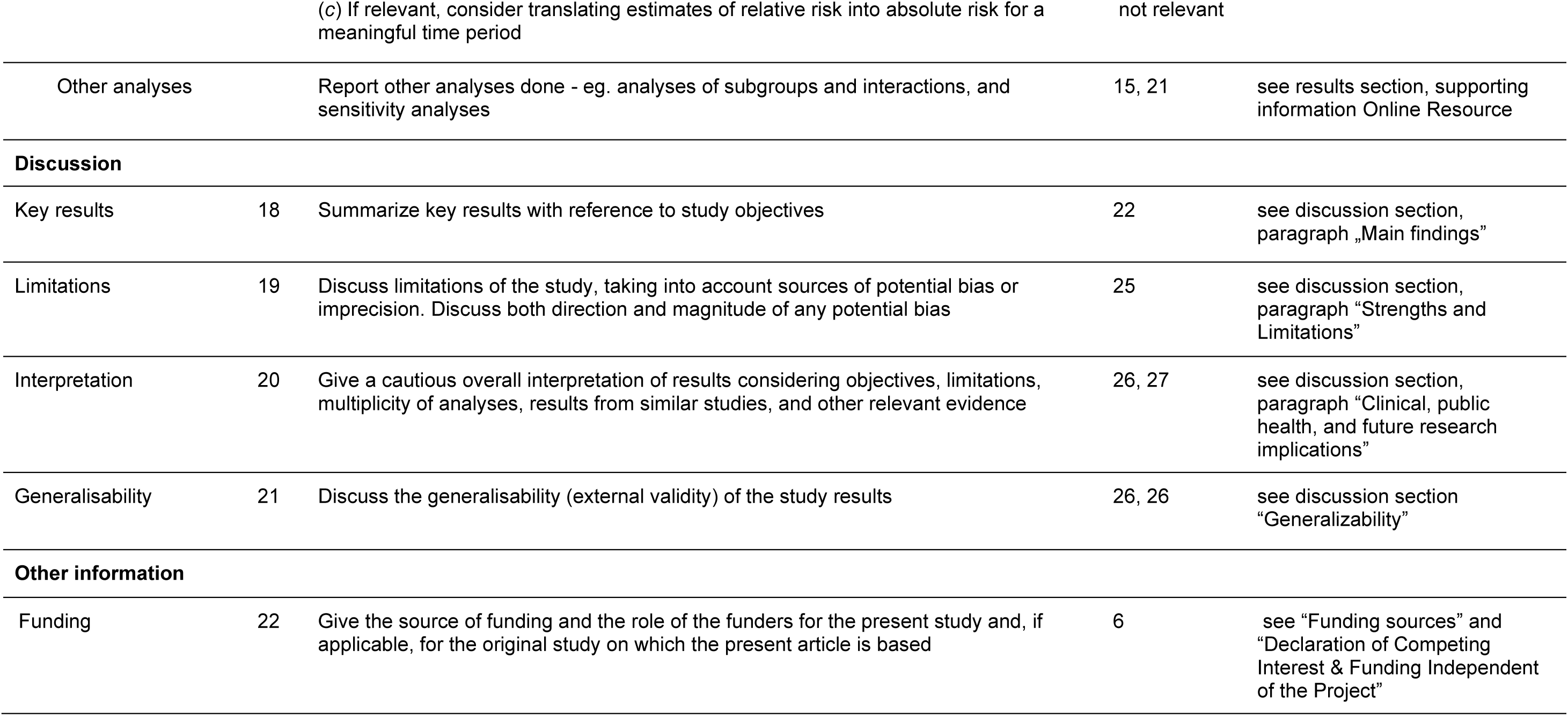

