## Supplementary material for "Mental distress and health-related quality of life among somatic inpatients: social gradients and hospital mental health implications based on the SomPsyNet study population": Online Resource 1 - Sample Comparison

**Online Resource 1. Comparison of the SomPsyNet Study Population with Hospital Registry Data, Regarding Patients' Gender and Age**

| | SomPsyNet Study Population | Hospital Registry Data | Expected Frequencies in SomPsyNet Sample Based on Hospital Registry Data | Chi2-Test: SomPsyNet vs. Hospital Registry |
|---|---|---|---|---|
| **Total number of patients** | **3,179** | **24,808** | **----** |  |
| **Gender** | | | | **8.20*10^-7** |
| male or other (No.) | 1,470 | 10,401 | 1,333 | |
| male or other (%) | 46.24% | 41.93% | | |
| female (No.) | 1,709 | 14,407 | 1,846 | |
| female (%) | 53.76% | 58.07% | | |
| **Age (in years)** | | | | **6.96*10^-49** |
| lower than 30 | 218 | 964 | 124 | |
| 30 – 39 | 308 | 1,750 | 224 | |
| 40 – 49 | 329 | 2,204 | 282 | |
| 50 – 59 | 483 | 3,362 | 431 | |
| 60 – 69 | 598 | 4,054 | 519 | |
| 70 – 79 | 678 | 5,689 | 729 | |
| at least 80 | 565 | 6,785 | 869 | |
