## Supplementary material for "Mental distress and health-related quality of life among somatic inpatients: social gradients and hospital mental health implications based on the SomPsyNet study population": Online Resource 2 - Sociodemographics

**Online Resource 2. Sociodemographic characteristics of the study sample**

| | Overall (N = 3,179) | | | |
|---|---|---|---|---|
| | N | % | lower 95%CI | upper 95%CI |
| **Age (in years)** | | | | |
| less than 30 | 218 | 6.86 | [2.91; | 15.32] |
| 30 - 39 | 308 | 9.69 | [3.72; | 22.94] |
| 40 - 49 | 329 | 10.35 | [5.53; | 18.53] |
| 50 - 59 | 483 | 15.19 | [10.81; | 20.94] |
| 60 - 69 | 598 | 18.81 | [12.95; | 26.51] |
| 70 - 79 | 678 | 21.33 | [14.49; | 30.25] |
| 80 and more | 565 | 17.77 | [8.62; | 33.13] |
| **Gender** | | | | |
| Female | 1,709 | 53.76 | [32.43; | 73.79] |
| Male or other | 1,470 | 46.24 | [26.21; | 67.57] |
| **Household income (in CHF per month)a** | | | | |
| less than 4,500 | 658 | 28.87 | [25.67; | 32.30] |
| 4,500 - 5,999 | 602 | 26.42 | [24.65; | 28.26] |
| 6,000 - 8,999 | 555 | 24.35 | [22.41; | 26.41] |
| 9,000 or more | 464 | 20.36 | [17.60; | 23.43] |
| **Nationality** | | | | |
| Swiss citizenship | 2,637 | 82.95 | [77.72; | 87.16] |
| No Swiss citizenship | 542 | 17.05 | [12.84; | 22.28] |
| **Marital statusb** | | | | |
| Divorced or dissolved partnership | 504 | 16.06 | [13.31; | 19.25] |
| Married or partnered | 1,470 | 46.85 | [40.14; | 53.67] |
| Single or unmarried | 785 | 25.02 | [16.76; | 35.60] |
| Widowed | 379 | 12.08 | [6.36; | 21.75] |
| **Living arrangmentc** | | | | |
| With spouse/partner/family/friends | 1,927 | 67.76 | [60.35; | 74.37] |
| Alone | 877 | 30.84 | [24.34; | 38.20] |
| In a nursing home / hospital or other long-term care facility / other | 40 | 1.41 | [0.84; | 2.33] |
| **Highest education leveld** | | | | |
| Did not attend school / primary level / not applicable / other | 119 | 3.80 | [2.38; | 5.99] |
| Lower secondary level I | 417 | 13.30 | [11.47; | 15.38] |
| Upper secondary level II | 1,467 | 46.79 | [42.15; | 51.50] |
| Tertiary level | 1,132 | 36.11 | [30.31; | 42.34] |
| **Employmente** | | | | |
| Employed or in training | 1,068 | 40.04 | [25.28; | 56.87] |
| Pension recipient | 1,199 | 44.96 | [28.80; | 62.26] |
| Homemaker | 127 | 4.76 | [3.70; | 6.12] |
| Disability / accident / physical illness / mental illness | 193 | 7.24 | [5.05; | 10.28] |
| Unemployed / looking for a job / in military service / other | 80 | 3.00 | [2.20; | 4.07] |

*[a] 900 missing values; [b] 41 missing values; [c] 335 missing values; [d] 44 missing values; [e] 512 missing values.*
*Abbreviations: CHF, Swiss Francs; CI, confidence interval.*
*Note. Categories with n < 10 are not shown for data protection reasons.*
