## Supplementary material for "Mental distress and health-related quality of life among somatic inpatients: social gradients and hospital mental health implications based on the SomPsyNet study population": Online Resource 3 - Supplemental Methods

### Online Resource 3 – Supplemental Material

#### Supplemental Methods

##### Detailed Statistical Methods

We performed descriptive statistical analyses, calculating frequencies, and measures of central tendency and dispersion, tailored to the scale and distribution of each variable. Where appropriate, we estimated 95% Confidence Intervals (CIs) using the margin of error formula based on the standard normal distribution (Z) for the observed proportion in a sample, accounting for the sample size and applying a continuity correction.

We conducted separate regression analyses for each outcome with the variables listed in Table 2 as predictors of (i) exceeding the established cut-off scores in the 8-item Patient Health Questionnaire (PHQ-8), the Generalized Anxiety Disorder-7 scale (GAD-7), the Somatic Symptom Disorder–B Criteria Scale (SSD-12), or any of these indicators (at least one above cut-off) and (ii) 'Mental Health Component Summary (MCS) score' of the Short Form (36) Health Survey (SF-36), Physical Health Component Summary (PCS) score of SF-36, European Quality of Life – 5 Dimensions questionnaire – 5 level version (EQ-5D-5L index), and Somatic Symptom Scale-8 (SSS-8) as continuous outcomes. Thereby, each predictor's association with the outcome was adjusted for the other predictors in the model. We estimated how age, gender, nationality, household income, marital status, living arrangement, highest educational level, employment situation, assessed at baseline, were associated with the outcomes. We performed tests for trends for the ordinal data, age, household income, and highest educational level to identify linear patterns or significant trends within these variables with ordered categories.

Prior to analysis, missing data in key predictor variables – marital status, living arrangement, household income, education level, and employment situation – were addressed. We evaluated the missing data mechanism to select an appropriate imputation method. Little's test for Missing Completely at Random (MCAR) indicated that the data were not MCAR ($p<0.05$). To assess the plausibility of the Missing at Random (MAR) assumption, we generated binary indicators of missingness and conducted logistic regression analyses, regressing these indicators on observed covariates (e.g., age, gender, nationality, and health outcomes). Significant associations between missingness and these variables suggested that the MAR assumption was reasonable. Consequently, Multiple Imputation by Chained Equations (MICE) was implemented under the MAR assumption to impute missing values. We implemented MICE via the Stata 'mi' framework. This approach allowed for the generation of multiple datasets, effectively encapsulating the inherent uncertainty in imputation. Thereby, we used multinomial logistic regression for nominal variables and ordinal logistic regression for ordinal variables. We created 100 imputed datasets and ensured the reproducibility of our analysis through a fixed seed. The data storage format for multiple imputed datasets was “mlong”. In this format, each patient is recorded multiple times once for each imputation.

We conducted logistic regression analyses for binary outcomes, with all sociodemographic predictors included simultaneously to adjust for confounding effects. Multicollinearity was assessed through Spearman’s correlation and Variance Inflation Factors (VIFs), with a maximum correlation coefficient of 0.44 and all VIFs below 5, indicating no significant multicollinearity concerns. For continuous outcomes (MCS, PCS, SSS-8, and EQ-5D-5L), we initially considered linear regression but rigorously evaluated linear regression assumptions. Residuals from initial models showed significant heteroskedasticity (Breusch-Pagan/Cook-Weisberg test, $p<0.001$) and deviations from normality (Shapiro-Wilk test, $p<0.001$), though visual assessments suggested approximate normality. Due to these violations, we employed generalized linear models (GLMs) as alternatives to improve model fit. Thereby, GLMs were tailored to each continuous outcome's characteristics as follows: MCS and PCS scores, scaled to range from 0 to 1, were modeled using Gaussian GLMs with log link function to accommodate scaling and heteroskedasticity. SSS-8 scores, exhibiting right-skew and overdispersion, were analyzed using a Gamma distribution with log link function. EQ-5D-5L values were scaled to range from 0 to 1 and modeled with a binomial distribution and probit link function. Model selection was based on Akaike Information Criterion (AIC) and Bayesian Information Criterion (BIC), with GLMs showing improved fit over linear models.

For clustering inherent in the survey design – originating from the multi-tiered data structure across three hospitals and their respective wards – we accounted for within-ward homogeneity and between-ward heterogeneity by applying Stata’s ‘svy’ framework, which was implemented as follows. The dataset encompassed information based on patients from three hospitals, with each hospital contributing patients from several selected wards that were further split to create a total of 21 distinct clusters. These 21 clusters, grouped at 7 cluster triplets, were the basis for the randomization in the larger context of the SomPsyNet stepped-wedge cluster randomized trial design, which however was not of further relevance for the here-presented analyses. Based on the 21 clusters, we created another clustering indicator with 9 categories, reflecting the 7 triplets of the 21 clusters as well as medical specialty differences of the clusters within the triplets. We implemented the statistical Stata ‘svy’ framework, using this 9-category clustering indicator and applying the Taylor series linearization method.
