## Supplementary material for "Mental distress and health-related quality of life among somatic inpatients: social gradients and hospital mental health implications based on the SomPsyNet study population": Online Resource 4 - Figure Flow Chart

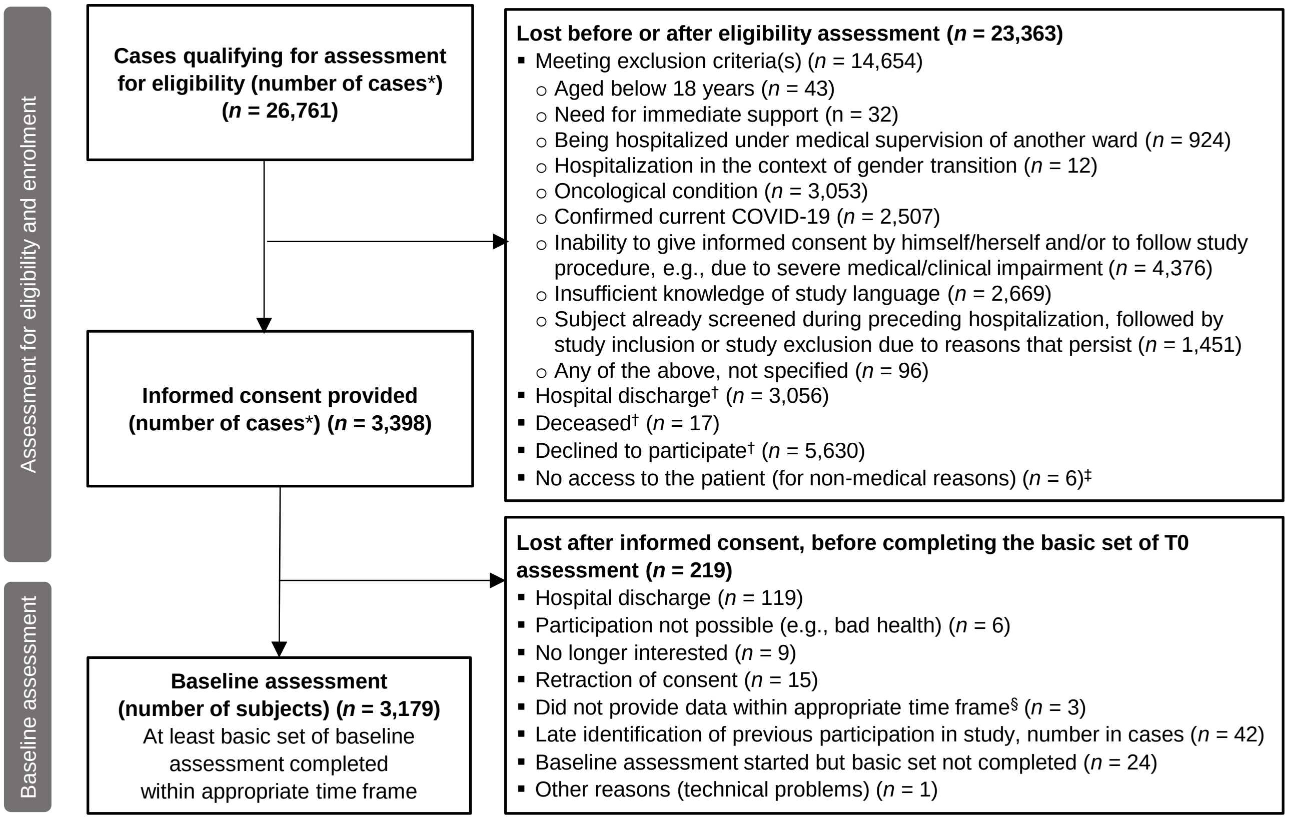


**Online Resource 4: Flowchart of the study sample**

**Due to multiple study centers, repeated recruitment and inclusion of the same patient could not always be prevented. Therefore, numbers are here shown in cases (i.e., the same subject could contribute to several cases).*
*†Not confirmed that we have no exclusion criteria.*
*‡Patients under police/security surveillance, who were not recruited for employee safety reasons.*
*§Completion of baseline assessment > 30 days after hospital discharge.*
*Abbreviations: COVID-19, Coronavirus disease 2019.*
