## Supplementary material for "Mental distress and health-related quality of life among somatic inpatients: social gradients and hospital mental health implications based on the SomPsyNet study population": Online Resource 5 - Results from Linear Regression

**Online Resource 5. Results from linear regression analyses, predicting the continuous variables MCS, PCS, SSS-8, and EQ-5D-5L by sociodemographic characteristics (N = 3,179)**

| | MCS[a] | | | PCS[b] | | | SSS-8[c] | | | EQ-5D-5L[d] | | |
|---|---|---|---|---|---|---|---|---|---|---|---|---|
| **Predictors** | Coefficients | lower 95%CI | upper 95%CI | Coefficients | lower 95%CI | upper 95%CI | Coefficients | lower 95%CI | upper 95%CI | Coefficients | lower 95%CI | upper 95%CI |
| **Age (in years)** | | | | | | | | | | | | |
| less than 30 | reference category | | | reference category | | | reference category | | | reference category | | |
| 30 - 39 | 2.37 | [-1.53; | 6.27] | -0.07 | [-2.73; | 2.59] | -1.16 | [-2.80; | 0.49] | 0.02 | [-0.07; | 0.11] |
| 40 - 49 | 2.13 | [-1.17; | 5.43] | -0.87 | [-5.12; | 3.38] | -1.17* | [-2.25; | -0.09] | 0.09* | [0.01; | 0.17] |
| 50 - 59 | 4.15 | [-0.28; | 8.59] | -4.63 | [-10.83; | 1.57] | -1.78* | [-3.32; | -0.25] | 0.07 | [-0.02; | 0.15] |
| 60 - 69 | 6.19* | [0.59; | 11.79] | -6.66 | [-15.58; | 2.26] | -2.67** | [-4.43; | -0.92] | 0.04 | [-0.06; | 0.14] |
| 70 - 79 | 7.07* | [1.05; | 13.09] | -9.42* | [-18.62; | -0.23] | -2.37* | [-3.98; | -0.76] | 0.01 | [-0.08; | 0.11] |
| 80 and more | 7.45* | [1.01; | 13.89] | -11.74* | [-19.88; | -3.60] | -2.57** | [-3.85; | -1.28] | 0.01 | [-0.07; | 0.09] |
| *Test for trend: F-value (dfs) p-value* | *8.37* | *(1,8)* | *0.02* | *9.22* | *(1,8)* | *0.02* | *22.08* | *(1,8)* | *<0.01* | *0.05* | *(1,8)* | *0.83* |
| **Gender** | | | | | | | | | | | | |
| Female | reference category | | | reference category | | | reference category | | | reference category | | |
| Male or other | -0.07 | [-2.65; | 2.52] | 0.76 | [-4.20; | 5.72] | -0.31 | [-1.64; | 1.03] | 0.07 | [-0.01; | 0.14] |
| **Household income (in CHF per month)** | | | | | | | | | | | | |
| Less than 4500 | reference category | | | reference category | | | reference category | | | reference category | | |
| 4500 - 5999 | 1.41 | [-0.78; | 3.61] | 1.07 | [-1.13; | 3.27] | -0.53 | [-1.37; | 0.31] | 0.03 | [-0.01; | 0.08] |
| 6000 - 8999 | 2.55* | [0.13; | 4.97] | 2.19 | [-0.48; | 4.86] | -0.72 | [-1.65; | 0.20] | 0.04* | [0; | 0.08] |
| 9000 or more | 4.35* | [1.01; | 7.70] | 2.26 | [-0.36; | 4.89] | -1.41* | [-2.51; | -0.31] | 0.06* | [0.02; | 0.11] |
| *Test for trend: F-value (dfs) p-value* | *11.30* | *(1,8)* | *<0.01* | *3.98* | *(1,8)* | *0.08* | *10.53* | *(1,8)* | *0.01* | *13.06* | *(1,8)* | *<0.01* |
| **Nationality** | | | | | | | | | | | | |
| Swiss citizenship | reference category | | | reference category | | | reference category | | | reference category | | |
| No Swiss citizenship | -1.94** | [-3.18; | -0.71] | -1.17 | [-2.97; | 0.64] | 0.97* | [0.27; | 1.66] | -0.02 | [-0.06; | 0.02] |
| **Marital status** | | | | | | | | | | | | |
| Divorced or dissolved partnership | -0.38 | [-2.65; | 1.89] | -0.10 | [-2.90; | 2.69] | 0.44 | [-0.28; | 1.16] | 0.03 | [-0.01; | 0.07] |
| Married or partnered | reference category | | | reference category | | | reference category | | | reference category | | |
| Single or unmarried | -0.07 | [-1.95; | 1.82] | -0.16 | [-3.64; | 3.32] | -0.12 | [-1.28; | 1.04] | 0.01 | [-0.07; | 0.08] |
| Widowed | 3.72** | [1.71; | 5.74] | -0.99 | [-4.03; | 2.06] | -0.86* | [-1.71; | -0.01] | 0.04 | [-0.01; | 0.10] |
| **Living arrangement** | | | | | | | | | | | | |
| With spouse/partner/family/friends | reference category | | | reference category | | | reference category | | | reference category | | |
| Alone | -0.60 | [-2.17; | 0.96] | -0.65 | [-3.08; | 1.77] | 0.08 | [-0.81; | 0.97] | -0.03 | [-0.07; | 0.01] |
| In a nursing home / hospital or other long-term care facility / other | -2.82 | [-8.70; | 3.06] | -6.04* | [-11.16; | -0.92] | 0.65 | [-1.20; | 2.51] | -0.21** | [-0.35; | -0.07] |
| **Highest educational level** | | | | | | | | | | | | |
| Did not attend school / primary level / not applicable / other | 0.91 | [-0.76; | 2.57] | 5.10* | [1.39; | 8.80] | -0.36 | [-1.55; | 0.83] | 0.10** | [0.03; | 0.17] |
| Lower secondary | 0.51 | [-3.10; | 4.13] | 2.73 | [-0.01; | 5.47] | 0.22 | [-0.75; | 1.20] | 0.09*** | [0.06; | 0.13] |
| Upper secondary | 0.77 | [-0.70; | 2.24] | 0.69 | [-1.60; | 2.99] | -0.03 | [-0.62; | 0.55] | 0.05** | [0.02; | 0.08] |
| Tertiary level | reference category | | | reference category | | | reference category | | | reference category | | |
| *Test for trend: F-value (dfs) p-value* | *1.65* | *(1,8)* | *0.24* | *14.20* | *(1,8)* | *<0.01* | *0.25* | *(1,8)* | *0.63* | *18.24* | *(1,8)* | *<0.01* |
| **Employment situation** | | | | | | | | | | | | |
| Employed or in training | reference category | | | reference category | | | reference category | | | reference category | | |
| Pension recipient | -1.45 | [-4.24; | 1.34] | -1.86 | [-4.38; | 0.66] | 0.56 | [-0.33; | 1.45] | 0.00 | [-0.05; | 0.05] |
| Homemaker | -3.51* | [-7.01; | -0.01] | -1.84 | [-6.83; | 3.15] | 1.20 | [-0.28; | 2.68] | -0.04 | [-0.10; | 0.02] |
| Disability / accident / physical illness / mental illness | -7.02** | [-9.96; | -4.07] | -9.10*** | [-11.95; | -6.24] | 2.45** | [1.40; | 3.51] | -0.14* | [-0.25; | -0.04] |
| Unemployed / looking for a job / in military service / other | -2.12 | [-5.50; | 1.26] | -2.54 | [-6.30; | 1.22] | -0.10 | [-1.71; | 1.51] | -0.05 | [-0.12; | 0.02] |

*[a] MCS-T-scores – 50 missing values; [b] PCS-T-scores 20 missing values; [c] Raw scores – 36 missing values; [d] Raw scores – 29 missing values.*
** p < 0.05; ** p < 0.01; *** p < 0.001.*
*Notes: Analyses conducted taking into account the sampling design and applying multiple imputation to handle missing data.*
*Abbreviations: CHF, Swiss Francs; CI, confidence interval; EQ-5D, European Quality of Life-5 Dimensions questionnaire; MCS, Mental Component Summary of the SF-36; PCS, Physical component summary of the SF-36; SSS-8, Somatic Symptom Scale-8.*
